# Patterns of brain degeneration in early-stage relapsing-remitting multiple sclerosis

**DOI:** 10.1101/2021.11.18.21266506

**Authors:** Rozanna Meijboom, Elizabeth N York, Agniete Kampaite, Mathew A Harris, Nicole White, Maria del C Valdés Hernández, Michael J Thrippleton, Peter Connick, David Hunt, Siddharthan Chandran, Adam D Waldman, on behalf of the FutureMS Consortium

## Abstract

Recurrent neuroinflammation in relapsing-remitting MS (RRMS) is thought to lead to neurodegeneration, resulting in progressive disability. Repeated magnetic resonance imaging (MRI) of the brain provides non-invasive measures of atrophy over time, a key marker of neurodegeneration. This study investigates regional neurodegeneration of the brain in early-stage RRMS using volumetry and voxel-based morphometry (VBM).

RRMS patients (N=354) underwent 3T structural MRI at diagnosis and 1-year follow-up, as part of the Scottish multicentre ‘FutureMS’ study. MRI data were processed using FreeSurfer to derive volumetrics, and FSL for VBM (grey matter (GM) only), to establish patterns of change in GM and normal-appearing white matter (NAWM) over time throughout the cerebrum, cerebellum and brainstem.

Volumetric analyses showed a decrease over time (q<0.05) in bilateral cortical GM and NAWM, multiple subcortical structures, cerebellar GM and the brainstem. Additionally, NAWM and GM volume decreased respectively in the following cortical regions, frontal: 14 out of 28 regions and 17/28; temporal: 18/18 and 15/18; parietal: 14/14 and 11/14; occipital: 7/8 and 8/8. Left GM and NAWM asymmetry was observed in the frontal lobe. GM VBM analysis showed three major clusters of decrease over time: 1) temporal lobe and subcortical areas, 2) cerebellum, 3) anterior cingulum and supplementary motor cortex; and four smaller clusters within the occipital lobe.

Widespread neurodegeneration was observed in early-stage RRMS; particularly in the brainstem, cerebellar GM, and subcortical and occipital-temporal regions. Volumetric and VBM results emphasise different as well as overlapping patterns of longitudinal change, and provide potential response markers for existing therapies and trials of neuroprotective agents.

## 1. Introduction

Multiple sclerosis (MS) is a neuroinflammatory and neurodegenerative disease affecting two million people worldwide [1–3]. MS prevalence varies geographically and is notably high in Scotland, with a prevalence of 290 cases per 100,000 population [4–6]. Disease progression and severity varies between individuals, and symptoms are diverse, including mobility and vision problems, pain, depression, fatigue and cognitive impairment [7]. Several MS subtypes have been identified, of which relapsing-remitting disease (RRMS) is the most common [8, 9]. There is currently no cure for MS and treatments targeting the neurodegenerative aspects of MS are limited. More accurate biomarkers of disease progression and improved understanding of disease mechanisms, particularly in terms of neurodegeneration, are required for more suitable treatment of MS. Magnetic resonance imaging (MRI) allows for studying neurodegeneration *in vivo* and may thus provide such valuable biomarkers of MS severity and progression.

White matter lesions (WMLs) on MRI, reflecting underlying inflammatory demyelinating lesions, are considered an imaging hallmark of RRMS and are required for RRMS diagnosis [10]. Modulating neuroinflammation is also the primary target for currently available disease modifying treatment (DMT) for RRMS. However, these appear to have only a limited effect on reducing associated neurodegenerative processes [11, 12]. Previous studies have shown that neurodegeneration also plays a prominent role in the disease evolution of RRMS and is importantly already present in the early stages of the disease [13–18]. Early-stage grey matter (GM) atrophy has been observed in specific brain areas, including the cingulate gyrus, precuneus, thalamus, basal ganglia, brainstem and cerebellum [14, 19–21]. Less regional detail appears to be known about WM atrophy in RRMS, but it also occurs in early stages and has been suggested to occur independently of WML development [13, 22, 23]. Importantly, atrophy appears to be a better predictor of clinical disability and deterioration than WMLs [16, 20, 24–34]. This suggests it could be an important target for disease-slowing treatments. There have indeed been studies investigating the effect of current DMTs on atrophy, suggesting a slowing of neurodegeneration after DMT use [35–40]. More research is required to investigate the effect of current DMTs and possible future neuroprotective therapies on brain atrophy. Such studies would benefit from detailed knowledge of the location and extent of early-stage GM and WM atrophy in RRMS [41].

To our knowledge, current literature does not provide detailed insights into regional WM volume loss in very early-stage RRMS, and only a limited number of studies have examined specific GM volume in early stages of the disease [14, 19–21]. The current study uses data from FutureMS [42], a large multicentre cohort of recently-diagnosed RRMS patients in Scotland, which acquired extensive MR imaging [43] at baseline and one-year follow-up. Importantly, study participants were recruited within six months of diagnosis, enabling early-stage detection of neurodegenerative patterns. In addition, participants were treatment-naive for the baseline study visit, improving detection of brain changes attributed to underlying MS disease mechanisms. Furthermore, FutureMS recruited people with RRMS throughout Scotland, allowing for studying brain degeneration in a cohort representative of the Scottish MS population. The aim of this study was to investigate comprehensive neurodegenerative changes in early-stage RRMS patients, supported by comparison of two different analysis approaches. This allows for identification of brain areas specifically affected by neurodegeneration, which may provide possible biomarkers of disease progression in terms of atrophy for DMT trials.

## 2. Methods

### 2.1 Participants

Participants with a recent diagnosis of RRMS according to the 2017 McDonald criteria (< 6 months) [10], were recruited across five neurology sites in Scotland: Aberdeen, Dundee, Edinburgh, Inverness and Glasgow, as part of the FutureMS study [42, 43]. Participants were 18 years or older and had the capacity to provide informed consent. Exclusion criteria were intake of DMTs prescribed prior to baseline assessment, participation in a clinical trial prior to baseline assessment and contraindications for MRI. Study visits took place at baseline (wave 0 [w0])) and 1-year follow-up (w1) and participants underwent brain MRI and expanded-disability status scale (EDSS) assessment at both time points. Full details of the FutureMS study have been previously described in Kearns et al. 2021 [42].

N=431 participants underwent MR imaging at w0 and N=382 participants underwent MR imaging at w1. The main reasons for not returning for w1 were not being able to reach participants at the provided contact details or participants having moved away from Scotland and not wanting to travel back. Additionally, the COVID-19 pandemic prematurely terminated w1 visits, which prevented several participants returning for their second visit. Additionally, 77/431 participants were excluded for various reasons (see Figure 1), resulting in N=354 available MRI datasets for analysis.

**Figure 1.**
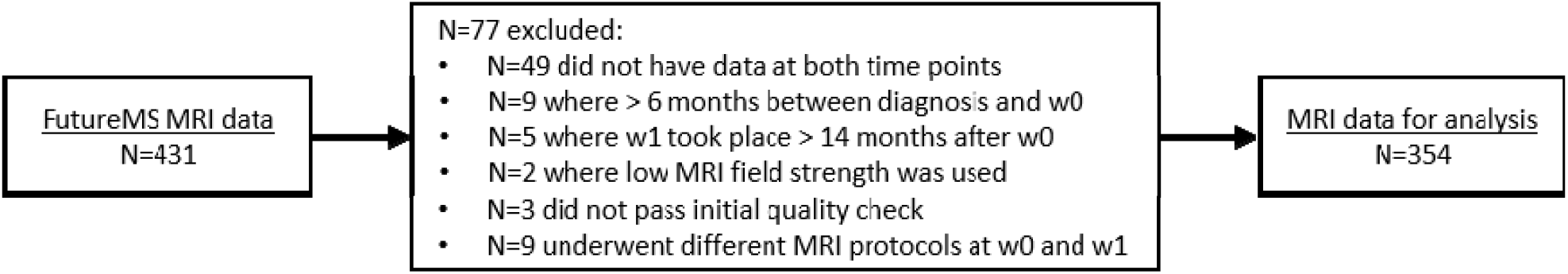
Overview of data exclusions.

All participants provided written informed consent before study entry. The study received ethical approval from the South East Scotland Research Ethics Committee 02 under reference 15/SS/0233 and was conducted in accordance with the Declaration of Helsinki and ICH guidelines on good clinical practice. All data were anonymised with unique study identifiers.

### 2.2 MR image acquisition

MR image acquisition was performed across four sites in Scotland using comparable 3T MRI systems (Siemens in Glasgow, Dundee and Edinburgh; Philips in Aberdeen). Protocol harmonisation was implemented during the course of the study to increase between-site comparability and facilitate image analysis. Importantly, each participant underwent both MRI scans at the same centre using the same protocol. All participants underwent T1-weighted, T2-weighted, and 2D and 3D FLAIR imaging. Full details have been previously described in Meijboom et al. (2021) [43] (see supplement Table 1 for an overview of all MR parameters).

### 2.3 MR image processing

Image processing methods have been previously described in full in Meijboom et al. (2021) [43] and are briefly summarised in the sections below.

#### 2.3.1 Registration and ICV

All images were registered to the T1W image with a rigid body transformation (degrees of freedom = 6) using FSL FLIRT (FSL v6.0.1) [44, 45] separately for each time point. Brain tissue was isolated using FSL BET2 (FSL v6.0.1) [46], followed by manual editing of the resulting w0 intracranial image using ITK-SNAP v3.8.0 [47]. For each participant, the edited w0 intracranial image was registered to the w1 T1W image, to avoid within-subject variability between time points. Intracranial volume (ICV) was extracted using fslstats (FSL6.0.1).

#### 2.3.2 WML segmentation

Hyperintense voxels on 2D FLAIR were identified by thresholding intensity values to 1.69 SDs > mean, using an adjusted method from Zhan et al. (2014) [48]. Resulting hyperintense areas unlikely to reflect pathology were removed using a pre-defined lesion distribution template [49], followed by Gaussian smoothing. Resulting WML masks were manually edited using ITK-SNAP [47]. WML volumes were extracted using fslstats and expressed as a ratio of ICV (r-ICV) to correct for head size.

#### 2.3.3 Tissue segmentation

Tissue segmentation was performed on T1W and T2W images using the longitudinal processing stream [50] in FreeSurfer v6.0 (http://surfer.nmr.mgh.harvard.edu/) with default settings, the Desikan-Kiliany atlas [51] and the edited ICV as brain mask. Freesurfer output was manually edited by a trained neuroscientist where necessary using FreeView v2.0 (FreeSurfer v6.0). WML masks were then subtracted from the resulting tissue segmentations using fslmaths (FSL6.0.1), to create final tissue masks. Global and regional tissue volumes (see Table 1) were calculated from the final tissue masks using fslstats, and subsequently expressed as r-ICV to correct for head size.

**Table 1.**
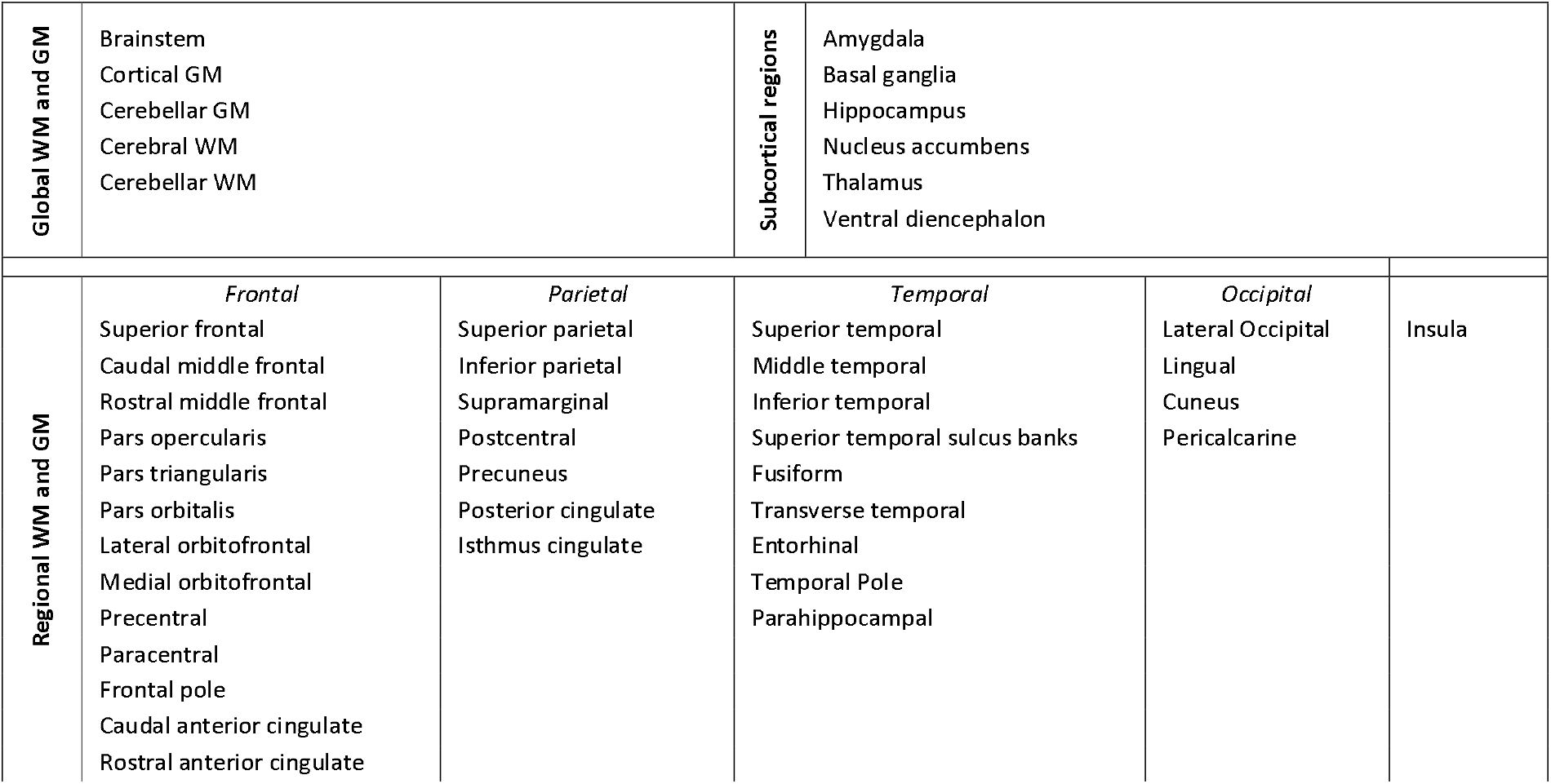
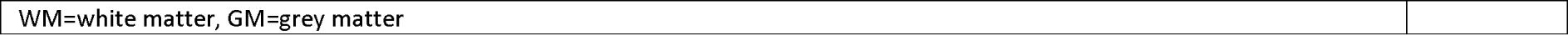
Global and regional GM/WM and subcortical areas. Left and right hemisphere were separately included for all areas, excluding the brainstem.

#### 2.3.4 Voxel-based morphometry

Longitudinal voxel-based morphometry (VBM), applying a voxel-wise comparison of local GM concentration between w0 and w1, was performed using the FSL VBM pipeline (FSL6.0.1; http://fsl.fmrib.ox.ac.uk/fsl/fslwiki/FSLVBM) [52–55]. It was performed for GM only, because GM VBM is a more commonly used and accepted method (e.g. a WM VBM pipeline is not implemented in FSL). The first step of the VBM pipeline (i.e. brain extraction) was omitted, as we were able to use the previously created intracranial brain images, as described above. In step two, GM, WM and CSF were segmented from T1W images and the study-specific GM template was created. Specifically, all subject GM images at both time points were affine-registered to the GM ICBM-152 template, and then concatenated and averaged to create a study-specific GM template. The subject GM images were then non-linearly registered to this template, and concatenated and averaged again to then create a final study-specific GM template in standard space (2×2×2mm^3^ resolution). In the last step, subject GM at both time points was then non-linearly registered to this study-specific template, after which a Jacobian modulation was applied, resulting in modulated GM images for each time point per subject.

As an additional step, to allow for longitudinal VBM analysis, we subtracted the modulated GM images for w1 from those at w0 using fslmaths, resulting in a GM difference file for each subject. Modulated GM difference files were then smoothed with a Gaussian size 2 kernel using fslmaths, and concatenated using fslmerge (FSL6.0.1) to form a final 4D group GM difference image for statistical analysis.

### 2.4 Statistical analyses

#### 2.4.1 Volumetrics

Statistical analysis for tissue volumes was performed using R v4.0.2 [56] and package lmerTest [57], with further packages ggplot2 [58] and lattice [59] used to create figures. A linear mixed-effects model was applied for each tissue volume (Table 1) with time point (w0, w1) as regressor of interest and age, sex, imaging site, WML change (w1 – w0) and DMT status at w1 as covariates. For each outcome variable, extreme outliers (>3 SD) as well as participants with missing data (for outcome and/or covariates) at either time point were excluded and subsequently scaled before being entered into the linear model. FDR correction was performed to adjust p-values for multiple comparisons, with corrected p-values (q-values) considered significant at q<0.05.

#### 2.4.2 Voxel-based morphometry

FSL Randomise (FSL v6.0.1) [60] was used to perform nonparametric permutation analysis on the VBM output. A general linear model (GLM) design matrix was created with GM difference, age, sex, imaging site, DMT status at w1 and subject-specific WML mask as explanatory variables (EV). EVs were recentered to zero where appropriate.

WMLs were included as EVs to ensure their exclusion from the GM images. Subject-specific WML EVs were created by a) registering all subject-specific WML masks at w0 and w1 to the subject’s modulated GM segmentation image at respectively w0 and w1, with the VBM transformation matrix that was created for registration of the native T1W to the group-specific template (fslmaths), and b) using FSL tool ‘setup_masks’ (FSL6.0.1; https://fsl.fmrib.ox.ac.uk/fsl/fslwiki/Randomise/UserGuide) to generate a concatenated 4D subject-specific WML image from the step a output images, as well as add a WML EV for each subject separately to the design matrix.

One t-contrast was defined, which assessed GM change over time, corrected for the remaining EV’s. The ‘randomise’ function was run using 5000 permutations, and with threshold-free cluster enhancing (TFCE) [61] to perform a voxel-wise analysis with a specific focus on voxel clusters. Cluster (FSL6.0.1) was used to extract resulting clusters and atlasquery (FSL v6.0.1) was used to obtain labels for the included anatomical regions. Results were reported for voxels at p<0.001 and family-wise error (FWR) corrected for multiple comparisons.

## 3. Results

### 3.1 Demographics

Data from N=315 participants were used for tissue volume analysis and data from N=351 were used for VBM analysis. Specifically, for tissue volume analysis 39/354 datasets were excluded after FreeSurfer output quality check and for VBM 3/354 datasets were excluded for data processing purposes. In addition, for the tissue volume dataset, for participants where segmentation failed for one or a small number of areas only (usually subcortical), the appropriate areas were excluded from analysis, but the remaining data of the participant was included in analysis. See Table 2 for participant demographics for each analysis type.

**Table 2.**
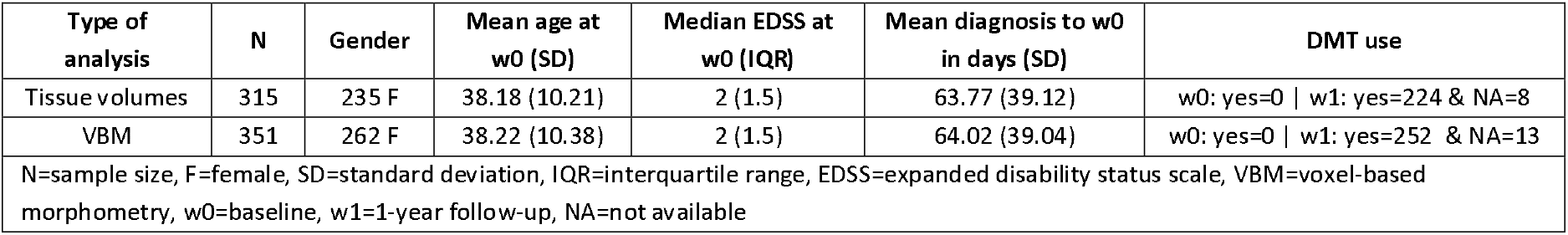
Participant demographics.

### 3.2 Global GM/WM, subcortical and whole-brain volumes

Whole-brain, brainstem and left and right cerebral GM and WM, cerebellar GM, brainstem, amygdala, basal ganglia, hippocampus, nucleus accumbens, thalamus and ventral diencephalon volumes significantly (q<0.05) decreased over time, whereas left and right cerebellar WM did *not* significantly (q>0.05) change over time (Table 3, Figure 2).

**Table 3.** Global GM/NAWM, subcortical and whole-brain volumes results for change over time (w1-w0) as assessed with a linear mixed-effects model, corrected for age, sex, imaging site, DMT status at w1 and WML change. Mean and SD are shown for raw volumes (r-ICV), without covariate correction.

|  | B | SE | df | t | p-value | CI 2.5. | CI 97.5. | Mean w0 | Mean w1 | SD w0 | SD w1 |
| --- | --- | --- | --- | --- | --- | --- | --- | --- | --- | --- | --- |
| Accumbens L | -0.0564 | 0.0213 | 306 | -2.6441 | **0.0086** | -0.0983 | -0.0145 | 0.0004 | 0.0004 | 0.0001 | 0.0001 |
| Accumbens R | -0.0797 | 0.0229 | 284 | -3.4809 | **0.0006** | -0.1246 | -0.0347 | 0.0004 | 0.0004 | 0.0001 | 0.0001 |
| Amygdala L | -0.1003 | 0.0228 | 305 | -4.3966 | **0.0000** | -0.1451 | -0.0555 | 0.0010 | 0.0010 | 0.0001 | 0.0001 |
| Amygdala R | -0.1484 | 0.0224 | 306 | -6.6139 | **0.0000** | -0.1924 | -0.1043 | 0.0012 | 0.0011 | 0.0001 | 0.0001 |
| Basal Ganglia L | -0.1545 | 0.0198 | 305 | -7.7884 | **0.0000** | -0.1934 | -0.1155 | 0.0065 | 0.0064 | 0.0008 | 0.0008 |
| Basal Ganglia R | -0.1632 | 0.0204 | 306 | -8.0073 | **0.0000** | -0.2032 | -0.1232 | 0.0065 | 0.0064 | 0.0008 | 0.0008 |
| Brainstem | -0.0654 | 0.0139 | 305 | -4.7223 | **0.0000** | -0.0926 | -0.0382 | 0.0132 | 0.0131 | 0.0014 | 0.0014 |
| GM cerebellar L | -0.0537 | 0.0119 | 305 | -4.5056 | **0.0000** | -0.0772 | -0.0303 | 0.0341 | 0.0339 | 0.0040 | 0.0039 |
| GM cerebellar R | -0.0624 | 0.0132 | 305 | -4.7386 | **0.0000** | -0.0882 | -0.0365 | 0.0346 | 0.0344 | 0.0039 | 0.0039 |
| GM cortical L | -0.0896 | 0.0172 | 305 | -5.1992 | **0.0000** | -0.1234 | -0.0558 | 0.1549 | 0.1541 | 0.0133 | 0.0132 |
| GM cortical R | -0.0626 | 0.0172 | 306 | -3.6346 | **0.0003** | -0.0964 | -0.0288 | 0.1551 | 0.1545 | 0.0131 | 0.0131 |
| Hippocampus L | -0.1067 | 0.0186 | 305 | -5.7223 | **0.0000** | -0.1433 | -0.0701 | 0.0027 | 0.0026 | 0.0003 | 0.0003 |
| Hippocampus R | -0.1122 | 0.0176 | 303 | -6.3582 | **0.0000** | -0.1468 | -0.0776 | 0.0028 | 0.0028 | 0.0007 | 0.0007 |
| NAWM cerebellar L | -0.0182 | 0.0171 | 304 | -1.0644 | 0.2880 | -0.0518 | 0.0154 | 0.0097 | 0.0097 | 0.0056 | 0.0054 |
| NAWM cerebellar R | 0.0091 | 0.0165 | 305 | 0.5507 | 0.5822 | -0.0233 | 0.0415 | 0.0091 | 0.0091 | 0.0012 | 0.0012 |
| NAWM cerebral L | -0.1394 | 0.0122 | 304 | -11.3883 | **0.0000** | -0.1635 | -0.1154 | 0.1403 | 0.1388 | 0.0137 | 0.0140 |
| NAWM cerebral R | -0.1274 | 0.0120 | 306 | -10.6280 | **0.0000** | -0.1509 | -0.1039 | 0.1402 | 0.1388 | 0.0135 | 0.0139 |
| Thalamus L | -0.1170 | 0.0118 | 306 | -9.9065 | **0.0000** | -0.1402 | -0.0938 | 0.0050 | 0.0049 | 0.0074 | 0.0073 |
| Thalamus R | -0.1058 | 0.0108 | 304 | -9.7489 | **0.0000** | -0.1271 | -0.0845 | 0.0044 | 0.0044 | 0.0005 | 0.0005 |
| Ventral DC L | -0.0794 | 0.0177 | 304 | -4.4856 | **0.0000** | -0.1142 | -0.0447 | 0.0026 | 0.0025 | 0.0003 | 0.0003 |
| Ventral DC R | -0.1328 | 0.0189 | 284 | -7.0208 | **0.0000** | -0.1699 | -0.0957 | 0.0024 | 0.0024 | 0.0007 | 0.0007 |
| Whole-brain | -0.1023 | 0.0150 | 305 | -6.8369 | **0.0000** | -0.1316 | -0.0729 | 0.7412 | 0.7374 | 0.0374 | 0.0378 |

B=standardised beta value, SE=standard error, df=degrees of freedom, CI=confidence interval for beta value, SD=standard deviation, w0=baseline, w1=1-year follow-up, L=left, R=right, GM=grey matter, NAWM=normal-appearing white matter, WML=white matter lesion, DC=diencephalon, r-ICV=ratio of intracranial volume. p-values surviving FDR correction (q<0.05) are highlighted in bold.

**Figure 2.**
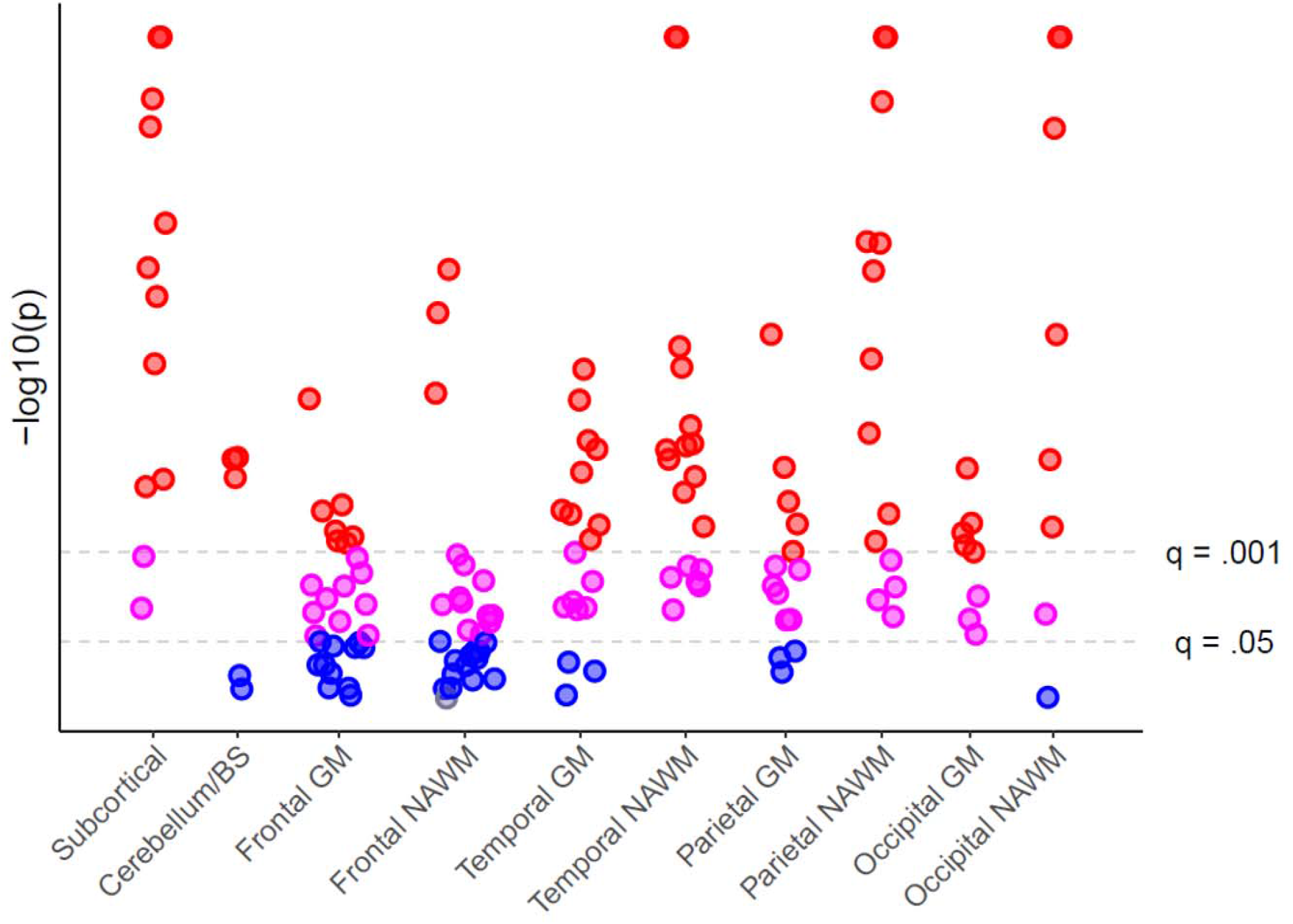
Manhattan plot of longitudinal brain volume change. Each point represents a tissue region within the given brain area category (x-axis). P-values were inverse log transformed (y-axis). Insignificant p-values (p>0.05) are shown in blue. Significant (p<0.05) FDR corrected p-values at q=0.05 are shown in pink, and in red for q=0.001. Regardless of significance, the direction of all effects – but one frontal NAWM area (in grey) - showed a longitudinal volume decrease. BS=brainstem, GM=grey matter, NAWM=normal-appearing white matter, FDR=false discovery rate.

**Figure 3.**
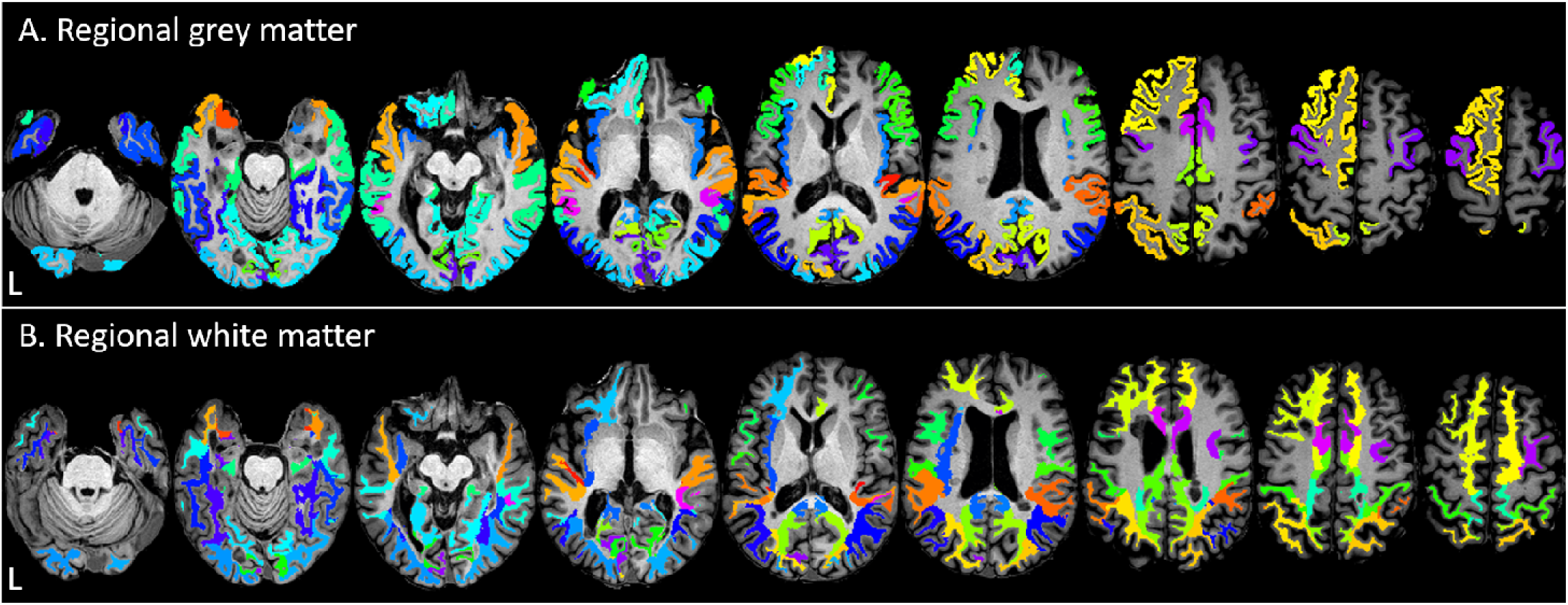
For illustration purposes, regional GM (A) and NAWM (B) regions with significant volume decrease (w1-w0; q<0.05) are shown on an example subject’s axial T1W image. Colours were chosen to emphasise borders between regions and have no further meaning in terms of results. This figure was created using MRIcron (https://www.nitrc.org/projects/mricron).

### 3.3 Regional cortical GM and cerebral WM

Regional GM and NAWM volume decrease (q<0.05) was observed in most regions and predominantly in the temporal, parietal and occipital lobe. GM volume decreased over time in 15/18 temporal regions, 15/26 frontal regions, 11/14 parietal regions, 8/8 occipital regions and the bilateral insula. NAWM volume decreased over time in 18/18 temporal regions, 13/26 frontal regions, 14/14 parietal regions, 7/8 occipital regions and the insula (L). More GM and NAWM regions showed volume loss in the left than in the right hemisphere, particularly in the frontal lobe. See Figures 2-3 and supplement Table 2.

### 3.4 Voxel-based GM change

Twelve clusters of various sizes (Table 5) showed a change in local concentration of GM over time. The largest clusters were centred within 1) the temporal lobe and subcortical areas, 2) cerebellum, 3) anterior cingulum and supplementary motor cortex. Additional smaller clusters were also observed in the temporal lobe and cerebellum. Furthermore, four smaller clusters were observed in the occipital lobe, as well as one cluster in the posterior frontal lobe and one in the parietal supramarginal and angular gyrus. See Figure 4.

**Table 5.** VBM cluster results (cluster-size >20, p_corrected_<0.001) for GM change over time (w1-w0) in RRMS (N=351).

| Cluster Index | Voxels | MAX X (mm) | MAX Y (mm) | MAX Z (mm) | COG X (mm) | COG Y (mm) | COG Z (mm) | MAX anatomical location | COG anatomical location |
| --- | --- | --- | --- | --- | --- | --- | --- | --- | --- |
| 1 | 7415 | 34 | 4 | -44 | 3.03 | -1.47 | -4.59 | Temporal Pole, Anterior Temporal Fusiform Gyrus, Anterior Inferior Temporal Gyrus | Right thalamus |
| 2 | 2882 | 20 | -64 | -52 | 1.76 | -65.2 | -32.1 | Cerebellum | Cerebellum |
| 3 | 794 | 0 | 22 | 20 | -0.111 | -1.98 | 45.5 | Anterior Cingulate Gyrus | Anterior Cingulate Gyrus, Supplementary Motor Cortex |
| 4 | 83 | 42 | 12 | 26 | 42.1 | 9.71 | 30.8 | Inferior/Middle Frontal Gyrus, Precentral Gyrus | Inferior/Middle Frontal Gyrus, Precentral Gyrus |
| 5 | 74 | -24 | -76 | -12 | -22.8 | -73.9 | -11.4 | Occipital Fusiform Gyrus, Lateral Occipital Gyrus, Lingual Gyrus | Occipital Fusiform Gyrus, Lingual Gyrus |
| 6 | 62 | 34 | -96 | -10 | 27.1 | -97.8 | -5.35 | Occipital Pole, lateral occipital cortex | Occipital Pole |
| 7 | 39 | 14 | -98 | 18 | 10.7 | -95.4 | 22.9 | Occipital Pole | Occipital Pole |
| 8 | 38 | 60 | -48 | 18 | 55.7 | -47.2 | 20.9 | Angular Gyrus, Posterior Supramarginal Gyrus | Angular Gyrus, Posterior Supramarginal Gyrus |
| 9 | 37 | -44 | -20 | -36 | -45.9 | -19.1 | -31.5 | Posterior Inferior Temporal Gyrus, Posterior Temporal Fusiform Gyrus | Posterior Inferior Temporal Gyrus, Posterior Temporal Fusiform Gyrus |
| 10 | 32 | 20 | -102 | 4 | 16.8 | -101 | 7.44 | Occipital Pole | Occipital Pole |
| 11 | 27 | -62 | -24 | 4 | -63.3 | -23.6 | 8.96 | Posterior Superior Temporal Gyrus, Planum Temporale | Posterior Superior Temporal Gyrus, Planum Temporale |
| 12 | 26 | -32 | -44 | -42 | -31.6 | -40.5 | -40.9 | Cerebellum | Cerebellum |
| VBM=voxel-based morphometry, GM=grey matter, RRMS=relapsing-remitting multiple sclerosis, MAX X/Y/Z=maximum cluster coordinates, mm=millimetres, COG X/Y/Z=centre of gravity cluster coordinates |  |  |  |  |  |  |  |  |  |

**Figure 4.**
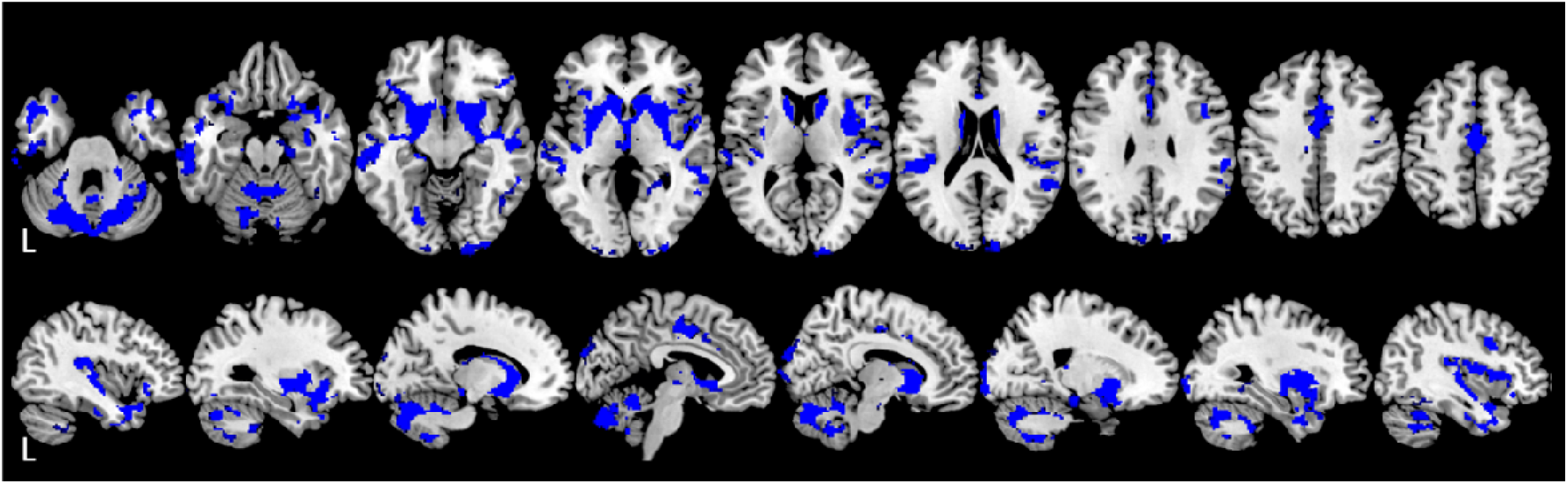
VBM results for significant GM change over time (w1-w0; p_corrected_<0.001; N=351) in RRMS are shown in blue on a template axial T1W image (top) and sagittal T1W image (bottom). This figure was created using MRIcron (https://www.nitrc.org/projects/mricron) and the implemented ‘ch2’ template (http://www.bic.mni.mcgill.ca/ServicesAtlases/Colin27).

## 4. Discussion

The current study used imaging data from the FutureMS cohort, a large longitudinal multicentre cohort of people with recently-diagnosed RRMS in Scotland. The aim of our study was to establish a profile of regional NAWM and GM volume change in the very early-stages of RRMS. In addition, the effects of different imaging analysis approaches were compared.

Widespread NAWM and GM atrophy was observed using both approaches. With the volumetric approach, early-stage RRMS showed brain volume loss in the brainstem, all subcortical regions and the cerebellar GM but not NAWM, over one year. Furthermore, cortical NAWM and GM volume loss over one year was also evident. Specifically, GM and NAWM volume decreased in nearly all temporal, parietal and occipital regions and in about half of frontal regions. Slightly more NAWM regions showed volume decrease in the temporal-parietal lobes and slightly more GM regions showed a volume decrease in the frontal lobe. Additionally, more *left* frontal GM and NAWM regions showed volume decrease than *right* frontal regions. With the VBM approach (GM only), most of these subcortical and cortical GM findings in early-stage RRMS were replicated. Large areas of change in GM concentration were observed in temporal and subcortical areas, as well as in the cerebellum and the anterior cingulum, with some smaller areas in the occipital lobe. However, overall, fewer regions of GM change were observed than with the volumetric approach, particularly in the parietal and frontal lobes, without prominent hemispheric differences.

### 4.1 Subcortical and brainstem atrophy

The importance of subcortical neurodegeneration in MS has been recently summarised by Ontaneda et al. (2021), emphasising that atrophy of deep GM structures is common in MS and may be a suitable target for DMT treatment [62]. In this study, we observed prominent subcortical volume change in early RRMS, which is in line with previous studies reporting early-stage subcortical atrophy, specifically in the thalamus and basal ganglia [14, 19, 24]. The thalamus has been suggested to play an important role in MS disease symptomatology, with lower volumes being predictive of clinical worsening [63], even in the early stages [64]. Basal ganglia changes have also frequently been observed and related to clinical change [65, 66].

Our study showed that other subcortical regions, such as the amygdala, hippocampus, ventral diencephalon and nucleus accumbens, also show early-stage atrophy. This is in line with previous studies reporting similar subcortical volume changes as well as associations with clinical changes in clinical isolated syndrome (CIS) or later stages of RRMS [66–73]. The brainstem is known to be involved in MS, showing both evidence of WML as well as volume loss, which are associated with clinical symptomatology [74–76]. Here we observed brainstem volume loss in a very early-stage of RRMS, which is corroborated by findings from Eshaghi et al. (2018) [14]. Overall, this may suggest that both the brainstem and subcortical regions play an important role in early-stage neurodegeneration in RRMS.

### 4.2 Cerebellar atrophy

The observed GM volume changes in this study are in line with previous studies reporting early-stage cerebellar atrophy [19] and they are unsurprising as the cerebellum has been shown to play an essential role in both sensorimotor and cognitive dysfunction in MS [77–80].

Cerebellar NAWM changes were not observed in this study, which is in line with previous studies reporting absence of cerebellar NAWM volume change in CIS and RRMS [81, 82]. In contrast, some studies do report cerebellar NAWM atrophy in MS [83, 84], however this may be explained by different disease stages being studied as well as study samples lacking a distinction between MS subtypes. Moreover, studies using diffusion tensor imaging (DTI) to explore cerebellar changes in RRMS have observed early-stage NAWM microstructural changes in absence of macrostructural NAWM volume change [85]. This may suggest that microstructural damage is already present in early-stage RRMS, which will go on to develop into macrostructural NAWM atrophy in later stages.

### 4.3 Cerebral regional NAWM and GM atrophy

This study suggests that both regional NAWM and GM are evident in early-stage RRMS in all brain lobes in variable degrees. Using volumetrics, cerebral GM volume loss was observed more prominently in the temporal, parietal and occipital lobes than in the frontal lobe. Similarly, with VBM, prominent GM changes were observed in the temporal, occipital and posterior frontal lobes, but less so in the anterior frontal lobe and parietal lobe. This discrepancy may be explained by the changes within the latter regions being less localised which may have left them undetected by VBM. The early GM changes observed here are partly in line with previous studies showing prominent early changes in the occipital [19], temporal [20, 73, 86], parietal (posterior cingulum) [14] and frontal lobe [86]. However, these observations differ from the current results in that they do not report on GM changes in all lobes simultaneously. This is likely explained by differences in methodology [87] (e.g. processing methods, inclusion of healthy controls, design), which is also supported by the discrepancy shown between VBM and volumetric results in this study.

Cerebral NAWM loss has been observed in previous studies [13, 22, 23], but to our knowledge, regional NAWM atrophy has not been investigated before. Similar to regional GM change, we observed widespread cerebral NAWM loss, independent of lesion accumulation, which was most prominent in the temporal, occipital and parietal regions. This suggests that NAWM neurodegeneration is also evident in early-stage RRMS. In comparison with cerebral GM volume loss, cerebral NAWM volume loss was observed in slightly more regions in temporal and parietal lobes, but not in the frontal lobe where more GM regions showed volume loss. This may support the notion that pathological processes underlying GM and NAWM change are at least partly dissociated [88].

More frontal NAWM and GM regions were affected in the left than in the right hemisphere - using the volumetric approach - whereas the temporal, parietal and occipital lobes were symmetrically affected. Although not specifically studied here, we can speculate that cognitive functions commonly associated with the left frontal lobe, such as language, may be more affected in early-stage MS than those associated with the right frontal lobe, such as non-verbal/spatial abilities [89]. Additionally, a left-hemispheric predilection for atrophy in all brain lobes has been observed previously in MS, however only by a small number of studies [90–92]. Furthermore, Preziosa et al (2017) [90] suggest that studies investigating hemispheric asymmetry in damage accumulation, in both ageing and neurological disease, have reported conflicting results. Overall, it remains unclear whether asymmetry of neurodegeneration in MS is evident and further research is required to elucidate this.

Surprisingly, we also observed a lack of volumetric changes within areas in the frontal lobe associated with sensorimotor functioning, such as in the precentral GM/NAWM and paracentral GM. However, paracentral NAWM volumetric change was indeed observed and the VBM results were also indicative of a small cluster of GM change within the supplemental motor area and precentral gyrus. Previous literature has shown GM volume change in the precentral gyrus in early RRMS compared with healthy controls [93], as well as abnormal iron deposition in the precentral gyrus in later stage MS [94]. This suggests that motor cortex abnormalities are involved in MS as expected, however the lack of volumetric precentral GM changes observed in our study may indicate that widespread motor cortex change over time may not be as fast as in other frontal regions in early RRMS.

### 4.4 Volumetry and VBM

Use of different imaging methods can lead to a difference in results, as was supported by the findings of this study. Although GM change was observed in overlapping areas as well here, there were key differences between the VBM and volumetric output. A volumetric approach like FreeSurfer segments brain regions, allowing for calculating the total amount of voxels within these regions. VBM on the other hand does not require regions to be pre-defined and looks for changes voxel-by-voxel, allowing for detection of local and possibly smaller changes [55]. Furthermore, FSL VBM also uses TFCE which enhances clusters of voxels that show change, increasing focus on changes that occur near each other, even when these changes are small [61]. Volumetrics on the other hand may not detect smaller changes, but it is more sensitive to distributed change across a region, which would be likely missed by VBM.

Further differences in processing steps and statistical methods are also likely to cause outcome differences. For example, VBM treats all data as one group and registers all participants to a common template, which may lead to some individual change being lost. FreeSurfer avoids this by processing participant data separately in their own native space. However, between-subject cortical variability can affect accuracy of atlas-based approaches and may thus bias the volumetric results [95]. Another example is that FSL VBM uses FWE correction for multiple comparisons, whereas we have applied FDR for the volumetric analysis. As FWE is considered to be more stringent than FDR, it is possible that VBM underestimated and FDR overestimated actual change, leading to a difference in results [96].

Specifically, for our RRMS cohort, the difference in results may indicate that the frontal and parietal changes observed with the volumetric approach are more widespread and more variable across participants. On the other hand, the overlapping temporal, occipital and subcortical results, may be indicative of prominent changes within these regions. Overall, both VBM and volumetric approaches have advantages and important limitations that should be taken into consideration when interpreting results.

### 4.5 Limitations

This study has some limitations. Firstly, FutureMS is a multi-centre study involving five different MR systems and two different (although very similar) MR protocols, which may have influenced the results. However, all participants underwent their MR assessment with the same system and protocol at both time points. Pooling of data across centres is important for clinical studies, as it allows for drawing conclusions based on larger datasets. Secondly, we have not studied the relationship between atrophy and clinical features in this study. The reason for this is that divergence in clinical disease course is limited over one year in early MS and will develop over a longer timescale. We are therefore initially concentrating on patterns of atrophy between the point of diagnosis and one year and will focus our future research on clinical correlates of atrophy.

### 4.6 Conclusion

Widespread neurodegeneration is observed in early-stage RRMS. This is observed particularly in the brainstem, cerebellar GM, subcortical regions and temporal-occipital GM and NAWM, as assessed by volumetry and VBM. Changes in these regions may provide a suitable target for DMT trials. Our future aims are directed at mapping neurodegenerative patterns across a ten-year time-period in the FutureMS cohort, as well as correlating regional atrophy with evolving clinical disability and other imaging and liquid biomarkers of neurodegeneration available in FutureMS.

## Data Availability

All data produced in the present study are available upon reasonable request to the authors.

## ACKNOWLEDGEMENTS

With thanks to FutureMS, hosted by Precision Medicine Scotland Innovation Centre (PMS-IC) and funded by a grant from the Chief Scientist Office, Scotland, to PMS-IC and Biogen Idec Ltd Insurance (combined funding under reference Exemplar SMS_IC010). We would like to thank other non-author contributors of the FutureMS consortium as follows: Mark Bastin, Chris Batchelor, Jessie Chang, Peter Connick, Fraser Brown, Tracy Brunton, Yingdi Chen, Shuna Colville, Annette Cooper, Rachel Dakin, Liz Elliott, David Hunt, Aidan Hutchison, Charlotte Jardine, Patrick Kearns, Lucy Kesseler, Michaela Kleynhans, Jen MacFarlane, Bev MacLennan, Sarah-Jane Martin, Daisy Mollison, Mary Monaghan, Scott Semple, Adam Scotson, Amy Stenson, Christine Weaver and Rosie Woodward. With special thanks to all FutureMS participants who have made this study possible.

Additional funding for authors came from the MS Society Edinburgh Centre for MS Research (grant reference 133; RM), Chief Scientist Office – SPRINT MND/MS program (ENY), NHS Lothian Research and Development Office (MJT), and the Row Fogo Charitable Trust (Grant No. BROD.FID3668413; MVH). Additional funding for the Edinburgh university 3T MRI Research scanner in RIE is funded by the Wellcome Trust (104916/Z/14/Z), Dunhill Trust (R380R/1114), Edinburgh and Lothians Health Foundation (2012/17), Muir Maxwell Research Fund, Edinburgh Imaging, and University of Edinburgh.

## SUPPLEMENT

**Supplement Table 1.** Future MS MRI parameters for protocol A and B

| A. PROTOCOL A |  |  |  |  |  |  |  |  |  |  |  |  |  |  |  |  |
| --- | --- | --- | --- | --- | --- | --- | --- | --- | --- | --- | --- | --- | --- | --- | --- | --- |
| Sequence |  | T1-weighted |  |  |  | T2-weighted |  |  |  | 2D FLAIR |  |  |  | 3D FLAIR |  |  |
| Site | EDI1 | GLA | DUN | ABN | EDI1 | GLA | DUN | ABN | EDI1 | GLA | DUN | ABN | EDI1 | GLA | DUN | ABN |
| Mode | 3D | 3D | 3D | 3D | 2D | 2D | 2D | 3D | 2D | 2D | 2D | 2D | 3D | 3D | 3D | 3D |
| FOV (mm) | 256 | 256 | 256 | 240 | 220 | 220 | 220 | 256 | 250 | 250 | 250 | 250 | 256 | 256 x 248 | 256 x 248 | 256 |
| Orientation | Sag | Sag | Sag | Sag | Ax | Ax | Ax | Sag | Ax | Ax | Ax | Ax | Sag | Sag | Sag | Sag |
| TR (ms) | 2530 | 2500 | 2500 | 3000 | 6000 | 6160 | 6160 | 2500 | 9500 | 9500 | 9500 | 11000 | 5000 | 5000 | 5000 | 8000 |
| TE (ms) | 3.37 | 2.26 | 2.26 | 3.9 | 96 | 96 | 96 | 310 | 124 | 124 | 124 | 125 | 715 | 393 | 393 | 347 |
| TI (ms) | 1100 | 1100 | 1100 | 1048 | - | - | - | - | 2400 | 2400 | 2400 | 2800 | 1800 | 1800 | 1800 | 2400 |
| Flip angle (deg) | 7 | 7 | 7 | 8 | 150 | 150 | 150 | - | 150 | 150 | 150 | 120 | - | - | - | - |
| Gap (mm) | - | - | - | - | 1.2 | 1.2 | 1.2 | - | 0 | 0 | 0 | 1 | - | - | - | - |
| Matrix (mm) | 256 x 256 | 256 x 256 | 256 x 256 | 240 x 240 | 320 x 320 | 320 x 314 | 314 x 314 | 256 x 256 | 256 x 256 | 256 x 256 | 256 x 256 | 252 x 226 | 256 x 256 | 256 x 248 | 256 x 248 | 256 x 256 |
| Voxel size (mm) | 1 x 1 x 1 | 1 x 1 x 1 | 1 x 1 x 1 | 1 x 1 x 1 | 0.7 x 0.7 x 4 | 0.7 x 0.7 x 4 | 0.7 x 0.7 x 4 | 1 x 1 x 2 | 1 x 1 x 3 | 1 x 1 x 3 | 1 x 1 x 3 | 1 x 1.1 x 3 | 1 x 1 x 1 | 1 x 1 x 1.3 | 1 x 1 x 1.3 | 1 x 1 x 2 |
| Slices reconstructed | 176 | 176 | 176 | 160 | 33 | 33 | 33 | 176 | 60 | 60 | 60 | 29 | 176 | 176 | 176 | 176 |
| Acq. Time (m:ss) | 6:03 | 5:59 | 5:59 | 5:38 | 1:26 | 1:03 | 1:03 | 3:42 | 7:38 | 7:38 | 7:38 | 5:08 | 7:22 | 5:32 | 5:32 | 8:32 |

| B. PROTOCOL B |  |  |  |  |  |  |  |  |  |  |  |  |  |  |
| --- | --- | --- | --- | --- | --- | --- | --- | --- | --- | --- | --- | --- | --- | --- |
| Sequence |  | T1-weighted (MPRAGE) |  |  |  | T2-weighted dual echo (FSE) |  |  |  | 2D FLAIR (PROPELLER) |  |  |  | 3D FLAIR (SPACE) |
| Mode |  | 3D |  |  |  | 2D |  |  |  | 2D |  |  |  | 3D |
| FOV (mm) |  | 256 |  |  |  | 250 |  |  |  | 250 |  |  |  | 256 |
| Orientation |  | Sagittal |  |  |  | Axial |  |  |  | Axial |  |  |  | Sagittal |
| TR (ms) |  | 2500 |  |  |  | 3630 |  |  |  | 9500 |  |  |  | 5000 |
| TE (ms) |  | 2.26 |  |  |  | 9.6, 96 |  |  |  | 120 |  |  |  | 393 |
| TI (ms) |  | 1100 |  |  |  | - |  |  |  | 2400 |  |  |  | 1800 |
| Flip angle (deg) |  | 7 |  |  |  | 150 |  |  |  | 150 |  |  |  | - |
| Gap (mm) |  | - |  |  |  | 0 |  |  |  | 0 |  |  |  | - |
| Matrix (mm) |  | 256 x 256 |  |  |  | 384 x 384 |  |  |  | 256 x 256 |  |  |  | 256 x 256 |
| Voxel size (mm) |  | 1 x 1 x 1 |  |  |  | 0.7 x 0.7 x 3 |  |  |  | 1 x 1 x 3 |  |  |  | 1 x 1 x 1 |
| Slices |  | 176 |  |  |  | 60 |  |  |  | 60 |  |  |  | 176 |
| Acceleration factor (in-plane x slice) |  | 2 x 1 |  |  |  | 3 x 1 |  |  |  | 2 x 1 |  |  |  | 2 x 1 |
| Acq. Time (m:ss) |  | 5:59 |  |  |  | 4:01 |  |  |  | 4:47 |  |  |  | 6:52 |

FLAIR = fluid attenuated inversion recovery; EDI1 = Edinburgh site 1; GLA = Glasgow; DUN = Dundee; ABN = Aberdeen; FOV = field of view; TR = repetition time; TE = echo time; TI = inversion time; deg = degree; acq. = acquisition; sag = sagittal; Ax = axial

**Supplement Table 2.**
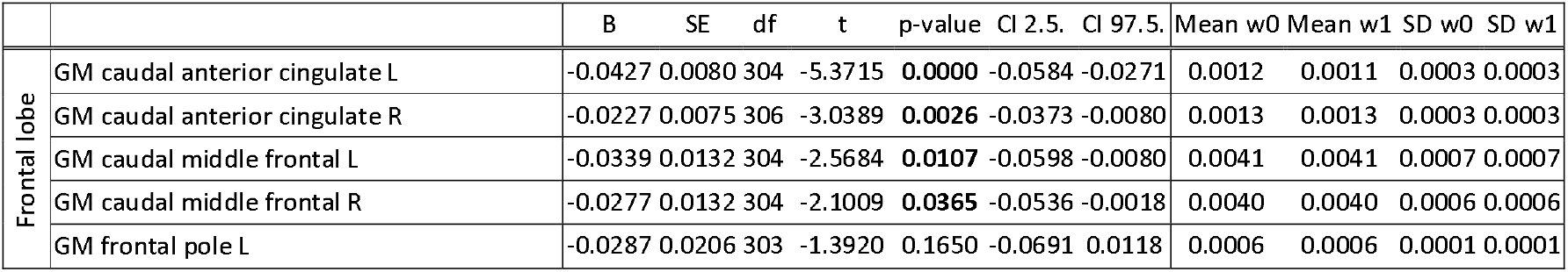
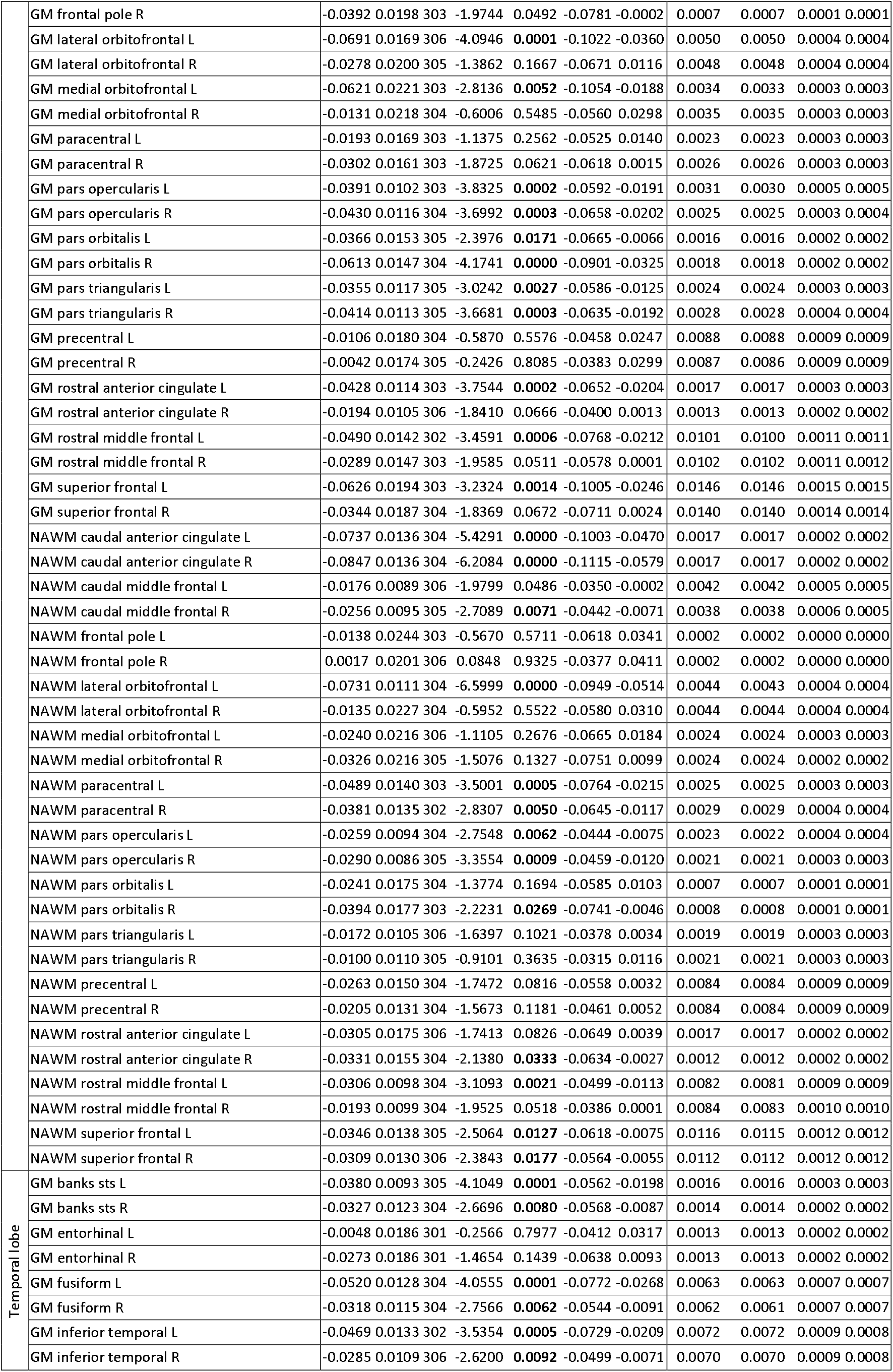
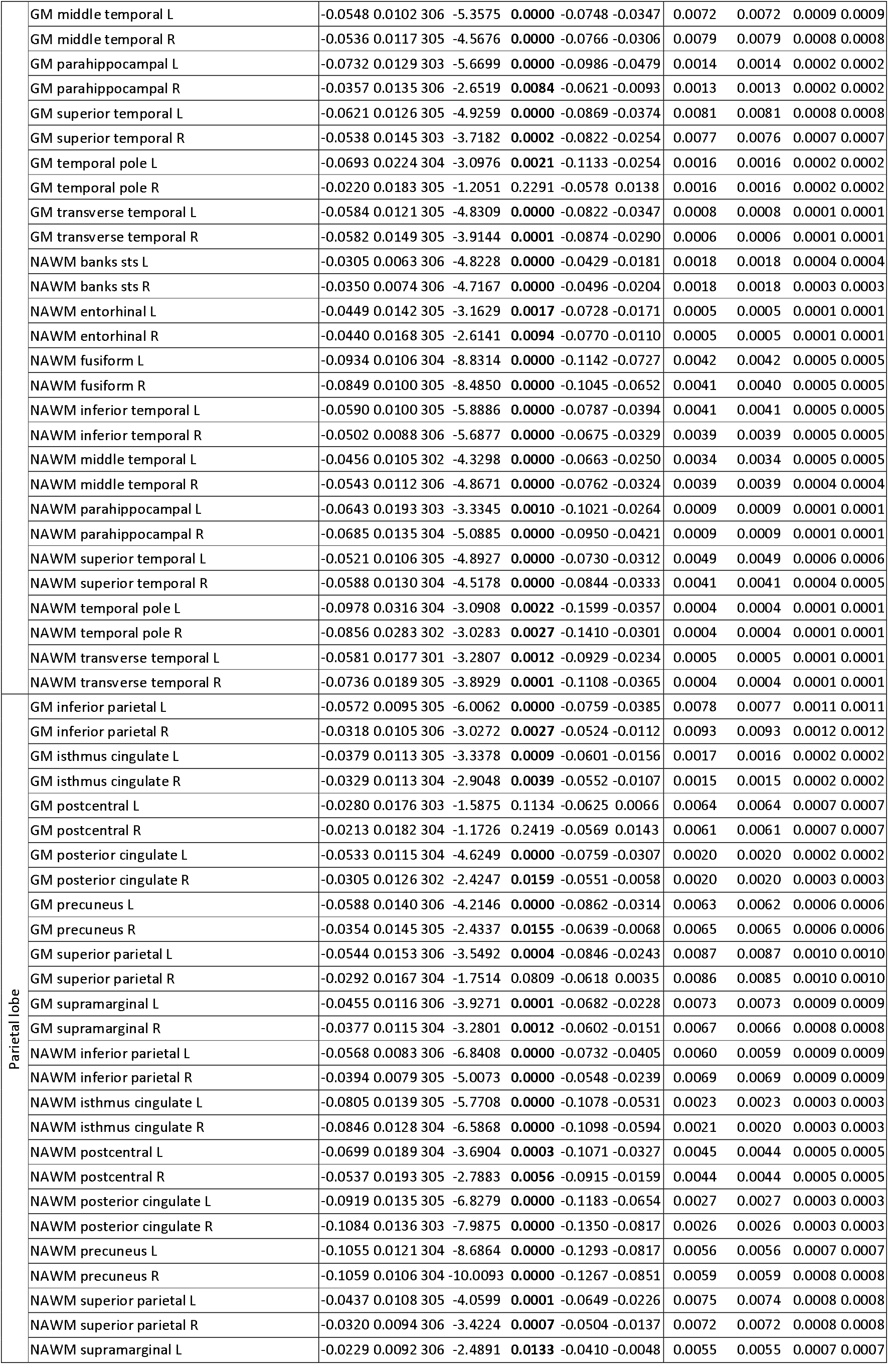
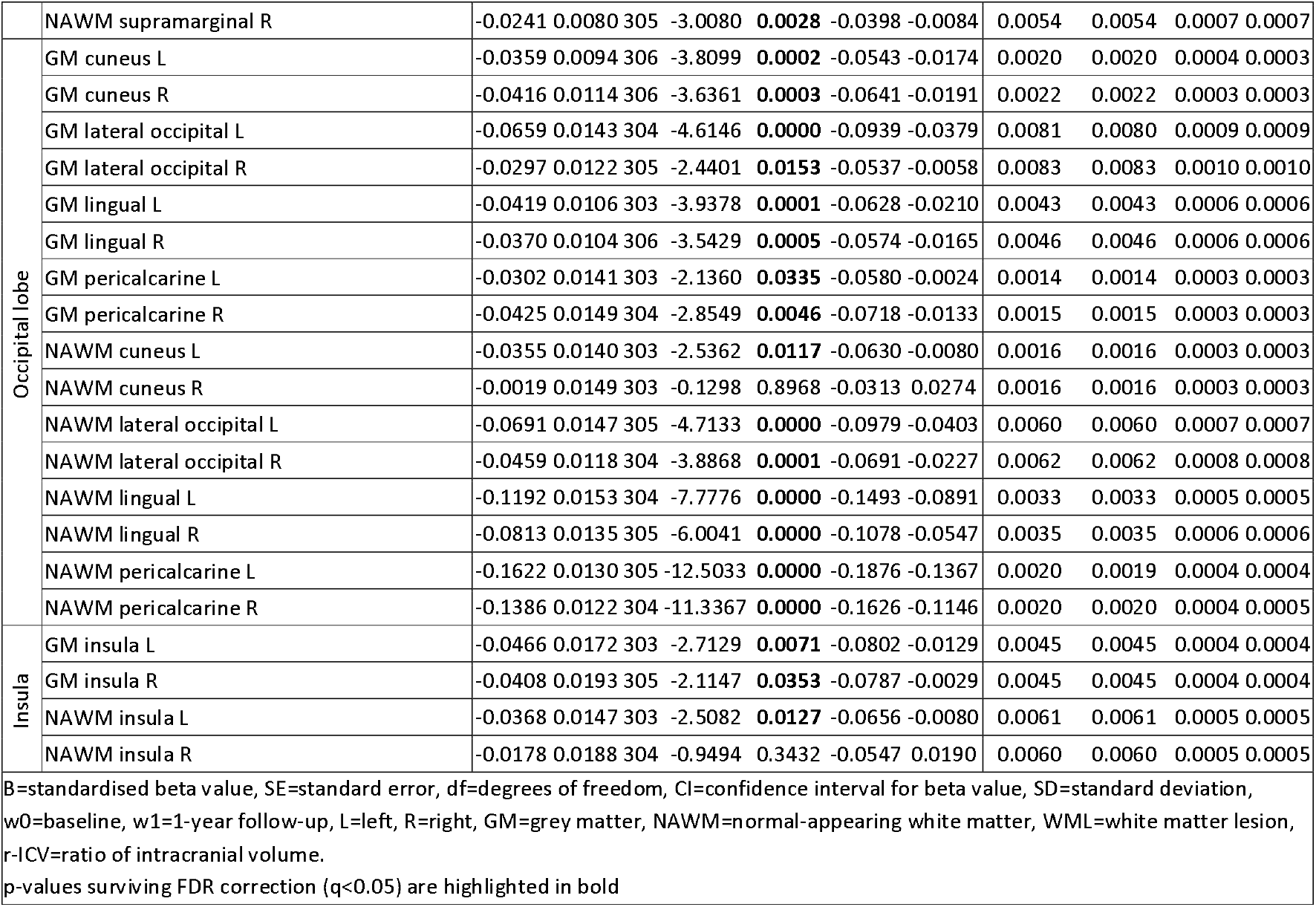
Regional GM and NAWM volume results for change over time (w1-w0) as assessed with a linear mixed-effects model, corrected for age, sex, imaging site, DMT status at w1 and WML change. Mean and SD are shown for raw volumes (r-ICV), without covariate correction.

