## Supplemental Table 1, Supplemental Table 2 for "Patterns of brain degeneration in early-stage relapsing-remitting multiple sclerosis" (from another version of this preprint: version 1, 2021-11-19)

SUPPLEMENT

**Supplement Table 1.** Future MS MRI parameters for protocol A and B

| A. PROTOCOL A | | | | | | | | | | | | | | | | |
| --- | --- | --- | --- | --- | --- | --- | --- | --- | --- | --- | --- | --- | --- | --- | --- | --- |
| Sequence | T1-weighted | | | | T2-weighted | | | | 2D FLAIR | | | | 3D FLAIR | | | |
| Site | **EDI1** | **GLA** | **DUN** | **ABN** | **EDI1** | **GLA** | **DUN** | **ABN** | **EDI1** | **GLA** | **DUN** | **ABN** | **EDI1** | **GLA** | **DUN** | **ABN** |
| Mode | 3D | 3D | 3D | 3D | 2D | 2D | 2D | 3D | 2D | 2D | 2D | 2D | 3D | 3D | 3D | 3D |
| FOV (mm) | 256 | 256 | 256 | 240 | 220 | 220 | 220 | 256 | 250 | 250 | 250 | 250 | 256 | 256 x 248 | 256 x 248 | 256 |
| Orientation | Sag | Sag | Sag | Sag | Ax | Ax | Ax | Sag | Ax | Ax | Ax | Ax | Sag | Sag | Sag | Sag |
| TR (ms) | 2530 | 2500 | 2500 | 3000 | 6000 | 6160 | 6160 | 2500 | 9500 | 9500 | 9500 | 11000 | 5000 | 5000 | 5000 | 8000 |
| TE (ms) | 3.37 | 2.26 | 2.26 | 3.9 | 96 | 96 | 96 | 310 | 124 | 124 | 124 | 125 | 715 | 393 | 393 | 347 |
| TI (ms) | 1100 | 1100 | 1100 | 1048 | - | - | - | - | 2400 | 2400 | 2400 | 2800 | 1800 | 1800 | 1800 | 2400 |
| Flip angle (deg) | 7 | 7 | 7 | 8 | 150 | 150 | 150 | - | 150 | 150 | 150 | 120 | - | - | - | - |
| Gap (mm) | - | - | - | - | 1.2 | 1.2 | 1.2 | - | 0 | 0 | 0 | 1 | - | - | - | - |
| Matrix (mm) | 256 × 256 | 256 × 256 | 256 × 256 | 240 × 240 | 320 x 320 | 320 × 314 | 314 x 314 | 256 x 256 | 256 x 256 | 256 × 256 | 256 x 256 | 252 x 226 | 256 x 256 | 256 x 248 | 256 x 248 | 256 x 256 |
| Voxel size (mm) | 1 × 1 × 1 | 1 × 1 × 1 | 1 × 1 × 1 | 1 x 1 x 1 | 0.7 × 0.7 × 4 | 0.7 × 0.7 × 4 | 0.7 × 0.7 × 4 | 1 x 1 x 2 | 1 x 1 x 3 | 1 × 1 × 3 | 1 x 1 x 3 | 1 x 1.1 x 3 | 1 x 1 x 1 | 1 x 1 x 1.3 | 1 x 1 x 1.3 | 1 x 1 x 2 |
| Slices reconstructed | 176 | 176 | 176 | 160 | 33 | 33 | 33 | 176 | 60 | 60 | 60 | 29 | 176 | 176 | 176 | 176 |
| Acq. Time (m:ss) | 6:03 | 5:59 | 5:59 | 5:38 | 1:26 | 1:03 | 1:03 | 3:42 | 7:38 | 7:38 | 7:38 | 5:08 | 7:22 | 5:32 | 5:32 | 8:32 |
| B. PROTOCOL B | | | | | | | | | | | | | | | | |
| Sequence | T1-weighted (MPRAGE) | | | | T2-weighted dual echo (FSE) | | | | 2D FLAIR (PROPELLER) | | | | 3D FLAIR (SPACE) | | | |
| Mode | 3D | | | | 2D | | | | 2D | | | | 3D | | | |
| FOV (mm) | 256 | | | | 250 | | | | 250 | | | | 256 | | | |
| Orientation | Sagittal | | | | Axial | | | | Axial | | | | Sagittal | | | |
| TR (ms) | 2500 | | | | 3630 | | | | 9500 | | | | 5000 | | | |
| TE (ms) | 2.26 | | | | 9.6, 96 | | | | 120 | | | | 393 | | | |
| TI (ms) | 1100 | | | | - | | | | 2400 | | | | 1800 | | | |
| Flip angle (deg) | 7 | | | | 150 | | | | 150 | | | | - | | | |
| Gap (mm) | - | | | | 0 | | | | 0 | | | | - | | | |
| Matrix (mm) | 256 x 256 | | | | 384 x 384 | | | | 256 x 256 | | | | 256 x 256 | | | |
| Voxel size (mm) | 1 x 1 x1 | | | | 0.7 x 0.7 x 3 | | | | 1 x 1 x 3 | | | | 1 x 1 x 1 | | | |
| Slices | 176 | | | | 60 | | | | 60 | | | | 176 | | | |
| Acceleration factor (in-plane × slice) | 2 × 1 | | | | 3 × 1 | | | | 2 × 1 | | | | 2 × 1 | | | |
| Acq. Time (m:ss) | 5:59 | | | | 4:01 | | | | 4:47 | | | | 6:52 | | | |
| FLAIR = fluid attenuated inversion recovery; EDI1 = Edinburgh site 1; GLA = Glasgow; DUN = Dundee; ABN = Aberdeen; FOV = field of view; TR = repetition time; TE = echo time; TI = inversion time; deg = degree; acq. = acquisition; sag = sagittal; Ax = axial | | | | | | | | | | | | | | | | |

**Supplement table 2**. Regional GM and NAWM volume results for change over time (w1-w0) as assessed with a linear mixed-effects model, corrected for age, sex, imaging site, DMT status at w1 and WML change. Mean and SD are shown for raw volumes (r-ICV), without covariate correction.

|  |  | B | SE | df | t | p-value | CI 2.5. | CI 97.5. | Mean w0 | Mean w1 | SD w0 | SD w1 |
| --- | --- | --- | --- | --- | --- | --- | --- | --- | --- | --- | --- | --- |
| Frontal lobe | GM caudal anterior cingulate L | -0.0427 | 0.0080 | 304 | -5.3715 | **0.0000** | -0.0584 | -0.0271 | 0.0012 | 0.0011 | 0.0003 | 0.0003 |
|  | GM caudal anterior cingulate R | -0.0227 | 0.0075 | 306 | -3.0389 | **0.0026** | -0.0373 | -0.0080 | 0.0013 | 0.0013 | 0.0003 | 0.0003 |
|  | GM caudal middle frontal L | -0.0339 | 0.0132 | 304 | -2.5684 | **0.0107** | -0.0598 | -0.0080 | 0.0041 | 0.0041 | 0.0007 | 0.0007 |
|  | GM caudal middle frontal R | -0.0277 | 0.0132 | 304 | -2.1009 | **0.0365** | -0.0536 | -0.0018 | 0.0040 | 0.0040 | 0.0006 | 0.0006 |
|  | GM frontal pole L | -0.0287 | 0.0206 | 303 | -1.3920 | 0.1650 | -0.0691 | 0.0118 | 0.0006 | 0.0006 | 0.0001 | 0.0001 |
|  | GM frontal pole R | -0.0392 | 0.0198 | 303 | -1.9744 | 0.0492 | -0.0781 | -0.0002 | 0.0007 | 0.0007 | 0.0001 | 0.0001 |
|  | GM lateral orbitofrontal L | -0.0691 | 0.0169 | 306 | -4.0946 | **0.0001** | -0.1022 | -0.0360 | 0.0050 | 0.0050 | 0.0004 | 0.0004 |
|  | GM lateral orbitofrontal R | -0.0278 | 0.0200 | 305 | -1.3862 | 0.1667 | -0.0671 | 0.0116 | 0.0048 | 0.0048 | 0.0004 | 0.0004 |
|  | GM medial orbitofrontal L | -0.0621 | 0.0221 | 303 | -2.8136 | **0.0052** | -0.1054 | -0.0188 | 0.0034 | 0.0033 | 0.0003 | 0.0003 |
|  | GM medial orbitofrontal R | -0.0131 | 0.0218 | 304 | -0.6006 | 0.5485 | -0.0560 | 0.0298 | 0.0035 | 0.0035 | 0.0003 | 0.0003 |
|  | GM paracentral L | -0.0193 | 0.0169 | 303 | -1.1375 | 0.2562 | -0.0525 | 0.0140 | 0.0023 | 0.0023 | 0.0003 | 0.0003 |
|  | GM paracentral R | -0.0302 | 0.0161 | 303 | -1.8725 | 0.0621 | -0.0618 | 0.0015 | 0.0026 | 0.0026 | 0.0003 | 0.0003 |
|  | GM pars opercularis L | -0.0391 | 0.0102 | 303 | -3.8325 | **0.0002** | -0.0592 | -0.0191 | 0.0031 | 0.0030 | 0.0005 | 0.0005 |
|  | GM pars opercularis R | -0.0430 | 0.0116 | 304 | -3.6992 | **0.0003** | -0.0658 | -0.0202 | 0.0025 | 0.0025 | 0.0003 | 0.0004 |
|  | GM pars orbitalis L | -0.0366 | 0.0153 | 305 | -2.3976 | **0.0171** | -0.0665 | -0.0066 | 0.0016 | 0.0016 | 0.0002 | 0.0002 |
|  | GM pars orbitalis R | -0.0613 | 0.0147 | 304 | -4.1741 | **0.0000** | -0.0901 | -0.0325 | 0.0018 | 0.0018 | 0.0002 | 0.0002 |
|  | GM pars triangularis L | -0.0355 | 0.0117 | 305 | -3.0242 | **0.0027** | -0.0586 | -0.0125 | 0.0024 | 0.0024 | 0.0003 | 0.0003 |
|  | GM pars triangularis R | -0.0414 | 0.0113 | 305 | -3.6681 | **0.0003** | -0.0635 | -0.0192 | 0.0028 | 0.0028 | 0.0004 | 0.0004 |
|  | GM precentral L | -0.0106 | 0.0180 | 304 | -0.5870 | 0.5576 | -0.0458 | 0.0247 | 0.0088 | 0.0088 | 0.0009 | 0.0009 |
|  | GM precentral R | -0.0042 | 0.0174 | 305 | -0.2426 | 0.8085 | -0.0383 | 0.0299 | 0.0087 | 0.0086 | 0.0009 | 0.0009 |
|  | GM rostral anterior cingulate L | -0.0428 | 0.0114 | 303 | -3.7544 | **0.0002** | -0.0652 | -0.0204 | 0.0017 | 0.0017 | 0.0003 | 0.0003 |
|  | GM rostral anterior cingulate R | -0.0194 | 0.0105 | 306 | -1.8410 | 0.0666 | -0.0400 | 0.0013 | 0.0013 | 0.0013 | 0.0002 | 0.0002 |
|  | GM rostral middle frontal L | -0.0490 | 0.0142 | 302 | -3.4591 | **0.0006** | -0.0768 | -0.0212 | 0.0101 | 0.0100 | 0.0011 | 0.0011 |
|  | GM rostral middle frontal R | -0.0289 | 0.0147 | 303 | -1.9585 | 0.0511 | -0.0578 | 0.0001 | 0.0102 | 0.0102 | 0.0011 | 0.0012 |
|  | GM superior frontal L | -0.0626 | 0.0194 | 303 | -3.2324 | **0.0014** | -0.1005 | -0.0246 | 0.0146 | 0.0146 | 0.0015 | 0.0015 |
|  | GM superior frontal R | -0.0344 | 0.0187 | 304 | -1.8369 | 0.0672 | -0.0711 | 0.0024 | 0.0140 | 0.0140 | 0.0014 | 0.0014 |
|  | NAWM caudal anterior cingulate L | -0.0737 | 0.0136 | 304 | -5.4291 | **0.0000** | -0.1003 | -0.0470 | 0.0017 | 0.0017 | 0.0002 | 0.0002 |
|  | NAWM caudal anterior cingulate R | -0.0847 | 0.0136 | 304 | -6.2084 | **0.0000** | -0.1115 | -0.0579 | 0.0017 | 0.0017 | 0.0002 | 0.0002 |
|  | NAWM caudal middle frontal L | -0.0176 | 0.0089 | 306 | -1.9799 | 0.0486 | -0.0350 | -0.0002 | 0.0042 | 0.0042 | 0.0005 | 0.0005 |
|  | NAWM caudal middle frontal R | -0.0256 | 0.0095 | 305 | -2.7089 | **0.0071** | -0.0442 | -0.0071 | 0.0038 | 0.0038 | 0.0006 | 0.0005 |
|  | NAWM frontal pole L | -0.0138 | 0.0244 | 303 | -0.5670 | 0.5711 | -0.0618 | 0.0341 | 0.0002 | 0.0002 | 0.0000 | 0.0000 |
|  | NAWM frontal pole R | 0.0017 | 0.0201 | 306 | 0.0848 | 0.9325 | -0.0377 | 0.0411 | 0.0002 | 0.0002 | 0.0000 | 0.0000 |
|  | NAWM lateral orbitofrontal L | -0.0731 | 0.0111 | 304 | -6.5999 | **0.0000** | -0.0949 | -0.0514 | 0.0044 | 0.0043 | 0.0004 | 0.0004 |
|  | NAWM lateral orbitofrontal R | -0.0135 | 0.0227 | 304 | -0.5952 | 0.5522 | -0.0580 | 0.0310 | 0.0044 | 0.0044 | 0.0004 | 0.0004 |
|  | NAWM medial orbitofrontal L | -0.0240 | 0.0216 | 306 | -1.1105 | 0.2676 | -0.0665 | 0.0184 | 0.0024 | 0.0024 | 0.0003 | 0.0003 |
|  | NAWM medial orbitofrontal R | -0.0326 | 0.0216 | 305 | -1.5076 | 0.1327 | -0.0751 | 0.0099 | 0.0024 | 0.0024 | 0.0002 | 0.0002 |
|  | NAWM paracentral L | -0.0489 | 0.0140 | 303 | -3.5001 | **0.0005** | -0.0764 | -0.0215 | 0.0025 | 0.0025 | 0.0003 | 0.0003 |
|  | NAWM paracentral R | -0.0381 | 0.0135 | 302 | -2.8307 | **0.0050** | -0.0645 | -0.0117 | 0.0029 | 0.0029 | 0.0004 | 0.0004 |
|  | NAWM pars opercularis L | -0.0259 | 0.0094 | 304 | -2.7548 | **0.0062** | -0.0444 | -0.0075 | 0.0023 | 0.0022 | 0.0004 | 0.0004 |
|  | NAWM pars opercularis R | -0.0290 | 0.0086 | 305 | -3.3554 | **0.0009** | -0.0459 | -0.0120 | 0.0021 | 0.0021 | 0.0003 | 0.0003 |
|  | NAWM pars orbitalis L | -0.0241 | 0.0175 | 304 | -1.3774 | 0.1694 | -0.0585 | 0.0103 | 0.0007 | 0.0007 | 0.0001 | 0.0001 |
|  | NAWM pars orbitalis R | -0.0394 | 0.0177 | 303 | -2.2231 | **0.0269** | -0.0741 | -0.0046 | 0.0008 | 0.0008 | 0.0001 | 0.0001 |
|  | NAWM pars triangularis L | -0.0172 | 0.0105 | 306 | -1.6397 | 0.1021 | -0.0378 | 0.0034 | 0.0019 | 0.0019 | 0.0003 | 0.0003 |
|  | NAWM pars triangularis R | -0.0100 | 0.0110 | 305 | -0.9101 | 0.3635 | -0.0315 | 0.0116 | 0.0021 | 0.0021 | 0.0003 | 0.0003 |
|  | NAWM precentral L | -0.0263 | 0.0150 | 304 | -1.7472 | 0.0816 | -0.0558 | 0.0032 | 0.0084 | 0.0084 | 0.0009 | 0.0009 |
|  | NAWM precentral R | -0.0205 | 0.0131 | 304 | -1.5673 | 0.1181 | -0.0461 | 0.0052 | 0.0084 | 0.0084 | 0.0009 | 0.0009 |
|  | NAWM rostral anterior cingulate L | -0.0305 | 0.0175 | 306 | -1.7413 | 0.0826 | -0.0649 | 0.0039 | 0.0017 | 0.0017 | 0.0002 | 0.0002 |
|  | NAWM rostral anterior cingulate R | -0.0331 | 0.0155 | 304 | -2.1380 | **0.0333** | -0.0634 | -0.0027 | 0.0012 | 0.0012 | 0.0002 | 0.0002 |
|  | NAWM rostral middle frontal L | -0.0306 | 0.0098 | 304 | -3.1093 | **0.0021** | -0.0499 | -0.0113 | 0.0082 | 0.0081 | 0.0009 | 0.0009 |
|  | NAWM rostral middle frontal R | -0.0193 | 0.0099 | 304 | -1.9525 | 0.0518 | -0.0386 | 0.0001 | 0.0084 | 0.0083 | 0.0010 | 0.0010 |
|  | NAWM superior frontal L | -0.0346 | 0.0138 | 305 | -2.5064 | **0.0127** | -0.0618 | -0.0075 | 0.0116 | 0.0115 | 0.0012 | 0.0012 |
|  | NAWM superior frontal R | -0.0309 | 0.0130 | 306 | -2.3843 | **0.0177** | -0.0564 | -0.0055 | 0.0112 | 0.0112 | 0.0012 | 0.0012 |
| Temporal lobe | GM banks sts L | -0.0380 | 0.0093 | 305 | -4.1049 | **0.0001** | -0.0562 | -0.0198 | 0.0016 | 0.0016 | 0.0003 | 0.0003 |
|  | GM banks sts R | -0.0327 | 0.0123 | 304 | -2.6696 | **0.0080** | -0.0568 | -0.0087 | 0.0014 | 0.0014 | 0.0002 | 0.0002 |
|  | GM entorhinal L | -0.0048 | 0.0186 | 301 | -0.2566 | 0.7977 | -0.0412 | 0.0317 | 0.0013 | 0.0013 | 0.0002 | 0.0002 |
|  | GM entorhinal R | -0.0273 | 0.0186 | 301 | -1.4654 | 0.1439 | -0.0638 | 0.0093 | 0.0013 | 0.0013 | 0.0002 | 0.0002 |
|  | GM fusiform L | -0.0520 | 0.0128 | 304 | -4.0555 | **0.0001** | -0.0772 | -0.0268 | 0.0063 | 0.0063 | 0.0007 | 0.0007 |
|  | GM fusiform R | -0.0318 | 0.0115 | 304 | -2.7566 | **0.0062** | -0.0544 | -0.0091 | 0.0062 | 0.0061 | 0.0007 | 0.0007 |
|  | GM inferior temporal L | -0.0469 | 0.0133 | 302 | -3.5354 | **0.0005** | -0.0729 | -0.0209 | 0.0072 | 0.0072 | 0.0009 | 0.0008 |
|  | GM inferior temporal R | -0.0285 | 0.0109 | 306 | -2.6200 | **0.0092** | -0.0499 | -0.0071 | 0.0070 | 0.0070 | 0.0009 | 0.0008 |
|  | GM middle temporal L | -0.0548 | 0.0102 | 306 | -5.3575 | **0.0000** | -0.0748 | -0.0347 | 0.0072 | 0.0072 | 0.0009 | 0.0009 |
|  | GM middle temporal R | -0.0536 | 0.0117 | 305 | -4.5676 | **0.0000** | -0.0766 | -0.0306 | 0.0079 | 0.0079 | 0.0008 | 0.0008 |
|  | GM parahippocampal L | -0.0732 | 0.0129 | 303 | -5.6699 | **0.0000** | -0.0986 | -0.0479 | 0.0014 | 0.0014 | 0.0002 | 0.0002 |
|  | GM parahippocampal R | -0.0357 | 0.0135 | 306 | -2.6519 | **0.0084** | -0.0621 | -0.0093 | 0.0013 | 0.0013 | 0.0002 | 0.0002 |
|  | GM superior temporal L | -0.0621 | 0.0126 | 305 | -4.9259 | **0.0000** | -0.0869 | -0.0374 | 0.0081 | 0.0081 | 0.0008 | 0.0008 |
|  | GM superior temporal R | -0.0538 | 0.0145 | 303 | -3.7182 | **0.0002** | -0.0822 | -0.0254 | 0.0077 | 0.0076 | 0.0007 | 0.0007 |
|  | GM temporal pole L | -0.0693 | 0.0224 | 304 | -3.0976 | **0.0021** | -0.1133 | -0.0254 | 0.0016 | 0.0016 | 0.0002 | 0.0002 |
|  | GM temporal pole R | -0.0220 | 0.0183 | 305 | -1.2051 | 0.2291 | -0.0578 | 0.0138 | 0.0016 | 0.0016 | 0.0002 | 0.0002 |
|  | GM transverse temporal L | -0.0584 | 0.0121 | 305 | -4.8309 | **0.0000** | -0.0822 | -0.0347 | 0.0008 | 0.0008 | 0.0001 | 0.0001 |
|  | GM transverse temporal R | -0.0582 | 0.0149 | 305 | -3.9144 | **0.0001** | -0.0874 | -0.0290 | 0.0006 | 0.0006 | 0.0001 | 0.0001 |
|  | NAWM banks sts L | -0.0305 | 0.0063 | 306 | -4.8228 | **0.0000** | -0.0429 | -0.0181 | 0.0018 | 0.0018 | 0.0004 | 0.0004 |
|  | NAWM banks sts R | -0.0350 | 0.0074 | 306 | -4.7167 | **0.0000** | -0.0496 | -0.0204 | 0.0018 | 0.0018 | 0.0003 | 0.0003 |
|  | NAWM entorhinal L | -0.0449 | 0.0142 | 305 | -3.1629 | **0.0017** | -0.0728 | -0.0171 | 0.0005 | 0.0005 | 0.0001 | 0.0001 |
|  | NAWM entorhinal R | -0.0440 | 0.0168 | 305 | -2.6141 | **0.0094** | -0.0770 | -0.0110 | 0.0005 | 0.0005 | 0.0001 | 0.0001 |
|  | NAWM fusiform L | -0.0934 | 0.0106 | 304 | -8.8314 | **0.0000** | -0.1142 | -0.0727 | 0.0042 | 0.0042 | 0.0005 | 0.0005 |
|  | NAWM fusiform R | -0.0849 | 0.0100 | 305 | -8.4850 | **0.0000** | -0.1045 | -0.0652 | 0.0041 | 0.0040 | 0.0005 | 0.0005 |
|  | NAWM inferior temporal L | -0.0590 | 0.0100 | 305 | -5.8886 | **0.0000** | -0.0787 | -0.0394 | 0.0041 | 0.0041 | 0.0005 | 0.0005 |
|  | NAWM inferior temporal R | -0.0502 | 0.0088 | 306 | -5.6877 | **0.0000** | -0.0675 | -0.0329 | 0.0039 | 0.0039 | 0.0005 | 0.0005 |
|  | NAWM middle temporal L | -0.0456 | 0.0105 | 302 | -4.3298 | **0.0000** | -0.0663 | -0.0250 | 0.0034 | 0.0034 | 0.0005 | 0.0005 |
|  | NAWM middle temporal R | -0.0543 | 0.0112 | 306 | -4.8671 | **0.0000** | -0.0762 | -0.0324 | 0.0039 | 0.0039 | 0.0004 | 0.0004 |
|  | NAWM parahippocampal L | -0.0643 | 0.0193 | 303 | -3.3345 | **0.0010** | -0.1021 | -0.0264 | 0.0009 | 0.0009 | 0.0001 | 0.0001 |
|  | NAWM parahippocampal R | -0.0685 | 0.0135 | 304 | -5.0885 | **0.0000** | -0.0950 | -0.0421 | 0.0009 | 0.0009 | 0.0001 | 0.0001 |
|  | NAWM superior temporal L | -0.0521 | 0.0106 | 305 | -4.8927 | **0.0000** | -0.0730 | -0.0312 | 0.0049 | 0.0049 | 0.0006 | 0.0006 |
|  | NAWM superior temporal R | -0.0588 | 0.0130 | 304 | -4.5178 | **0.0000** | -0.0844 | -0.0333 | 0.0041 | 0.0041 | 0.0004 | 0.0005 |
|  | NAWM temporal pole L | -0.0978 | 0.0316 | 304 | -3.0908 | **0.0022** | -0.1599 | -0.0357 | 0.0004 | 0.0004 | 0.0001 | 0.0001 |
|  | NAWM temporal pole R | -0.0856 | 0.0283 | 302 | -3.0283 | **0.0027** | -0.1410 | -0.0301 | 0.0004 | 0.0004 | 0.0001 | 0.0001 |
|  | NAWM transverse temporal L | -0.0581 | 0.0177 | 301 | -3.2807 | **0.0012** | -0.0929 | -0.0234 | 0.0005 | 0.0005 | 0.0001 | 0.0001 |
|  | NAWM transverse temporal R | -0.0736 | 0.0189 | 305 | -3.8929 | **0.0001** | -0.1108 | -0.0365 | 0.0004 | 0.0004 | 0.0001 | 0.0001 |
| Parietal lobe | GM inferior parietal L | -0.0572 | 0.0095 | 305 | -6.0062 | **0.0000** | -0.0759 | -0.0385 | 0.0078 | 0.0077 | 0.0011 | 0.0011 |
|  | GM inferior parietal R | -0.0318 | 0.0105 | 306 | -3.0272 | **0.0027** | -0.0524 | -0.0112 | 0.0093 | 0.0093 | 0.0012 | 0.0012 |
|  | GM isthmus cingulate L | -0.0379 | 0.0113 | 305 | -3.3378 | **0.0009** | -0.0601 | -0.0156 | 0.0017 | 0.0016 | 0.0002 | 0.0002 |
|  | GM isthmus cingulate R | -0.0329 | 0.0113 | 304 | -2.9048 | **0.0039** | -0.0552 | -0.0107 | 0.0015 | 0.0015 | 0.0002 | 0.0002 |
|  | GM postcentral L | -0.0280 | 0.0176 | 303 | -1.5875 | 0.1134 | -0.0625 | 0.0066 | 0.0064 | 0.0064 | 0.0007 | 0.0007 |
|  | GM postcentral R | -0.0213 | 0.0182 | 304 | -1.1726 | 0.2419 | -0.0569 | 0.0143 | 0.0061 | 0.0061 | 0.0007 | 0.0007 |
|  | GM posterior cingulate L | -0.0533 | 0.0115 | 304 | -4.6249 | **0.0000** | -0.0759 | -0.0307 | 0.0020 | 0.0020 | 0.0002 | 0.0002 |
|  | GM posterior cingulate R | -0.0305 | 0.0126 | 302 | -2.4247 | **0.0159** | -0.0551 | -0.0058 | 0.0020 | 0.0020 | 0.0003 | 0.0003 |
|  | GM precuneus L | -0.0588 | 0.0140 | 306 | -4.2146 | **0.0000** | -0.0862 | -0.0314 | 0.0063 | 0.0062 | 0.0006 | 0.0006 |
|  | GM precuneus R | -0.0354 | 0.0145 | 305 | -2.4337 | **0.0155** | -0.0639 | -0.0068 | 0.0065 | 0.0065 | 0.0006 | 0.0006 |
|  | GM superior parietal L | -0.0544 | 0.0153 | 306 | -3.5492 | **0.0004** | -0.0846 | -0.0243 | 0.0087 | 0.0087 | 0.0010 | 0.0010 |
|  | GM superior parietal R | -0.0292 | 0.0167 | 304 | -1.7514 | 0.0809 | -0.0618 | 0.0035 | 0.0086 | 0.0085 | 0.0010 | 0.0010 |
|  | GM supramarginal L | -0.0455 | 0.0116 | 306 | -3.9271 | **0.0001** | -0.0682 | -0.0228 | 0.0073 | 0.0073 | 0.0009 | 0.0009 |
|  | GM supramarginal R | -0.0377 | 0.0115 | 304 | -3.2801 | **0.0012** | -0.0602 | -0.0151 | 0.0067 | 0.0066 | 0.0008 | 0.0008 |
|  | NAWM inferior parietal L | -0.0568 | 0.0083 | 306 | -6.8408 | **0.0000** | -0.0732 | -0.0405 | 0.0060 | 0.0059 | 0.0009 | 0.0009 |
|  | NAWM inferior parietal R | -0.0394 | 0.0079 | 305 | -5.0073 | **0.0000** | -0.0548 | -0.0239 | 0.0069 | 0.0069 | 0.0009 | 0.0009 |
|  | NAWM isthmus cingulate L | -0.0805 | 0.0139 | 305 | -5.7708 | **0.0000** | -0.1078 | -0.0531 | 0.0023 | 0.0023 | 0.0003 | 0.0003 |
|  | NAWM isthmus cingulate R | -0.0846 | 0.0128 | 304 | -6.5868 | **0.0000** | -0.1098 | -0.0594 | 0.0021 | 0.0020 | 0.0003 | 0.0003 |
|  | NAWM postcentral L | -0.0699 | 0.0189 | 304 | -3.6904 | **0.0003** | -0.1071 | -0.0327 | 0.0045 | 0.0044 | 0.0005 | 0.0005 |
|  | NAWM postcentral R | -0.0537 | 0.0193 | 305 | -2.7883 | **0.0056** | -0.0915 | -0.0159 | 0.0044 | 0.0044 | 0.0005 | 0.0005 |
|  | NAWM posterior cingulate L | -0.0919 | 0.0135 | 305 | -6.8279 | **0.0000** | -0.1183 | -0.0654 | 0.0027 | 0.0027 | 0.0003 | 0.0003 |
|  | NAWM posterior cingulate R | -0.1084 | 0.0136 | 303 | -7.9875 | **0.0000** | -0.1350 | -0.0817 | 0.0026 | 0.0026 | 0.0003 | 0.0003 |
|  | NAWM precuneus L | -0.1055 | 0.0121 | 304 | -8.6864 | **0.0000** | -0.1293 | -0.0817 | 0.0056 | 0.0056 | 0.0007 | 0.0007 |
|  | NAWM precuneus R | -0.1059 | 0.0106 | 304 | -10.0093 | **0.0000** | -0.1267 | -0.0851 | 0.0059 | 0.0059 | 0.0008 | 0.0008 |
|  | NAWM superior parietal L | -0.0437 | 0.0108 | 305 | -4.0599 | **0.0001** | -0.0649 | -0.0226 | 0.0075 | 0.0074 | 0.0008 | 0.0008 |
|  | NAWM superior parietal R | -0.0320 | 0.0094 | 306 | -3.4224 | **0.0007** | -0.0504 | -0.0137 | 0.0072 | 0.0072 | 0.0008 | 0.0008 |
|  | NAWM supramarginal L | -0.0229 | 0.0092 | 306 | -2.4891 | **0.0133** | -0.0410 | -0.0048 | 0.0055 | 0.0055 | 0.0007 | 0.0007 |
|  | NAWM supramarginal R | -0.0241 | 0.0080 | 305 | -3.0080 | **0.0028** | -0.0398 | -0.0084 | 0.0054 | 0.0054 | 0.0007 | 0.0007 |
| Occipital lobe | GM cuneus L | -0.0359 | 0.0094 | 306 | -3.8099 | **0.0002** | -0.0543 | -0.0174 | 0.0020 | 0.0020 | 0.0004 | 0.0003 |
|  | GM cuneus R | -0.0416 | 0.0114 | 306 | -3.6361 | **0.0003** | -0.0641 | -0.0191 | 0.0022 | 0.0022 | 0.0003 | 0.0003 |
|  | GM lateral occipital L | -0.0659 | 0.0143 | 304 | -4.6146 | **0.0000** | -0.0939 | -0.0379 | 0.0081 | 0.0080 | 0.0009 | 0.0009 |
|  | GM lateral occipital R | -0.0297 | 0.0122 | 305 | -2.4401 | **0.0153** | -0.0537 | -0.0058 | 0.0083 | 0.0083 | 0.0010 | 0.0010 |
|  | GM lingual L | -0.0419 | 0.0106 | 303 | -3.9378 | **0.0001** | -0.0628 | -0.0210 | 0.0043 | 0.0043 | 0.0006 | 0.0006 |
|  | GM lingual R | -0.0370 | 0.0104 | 306 | -3.5429 | **0.0005** | -0.0574 | -0.0165 | 0.0046 | 0.0046 | 0.0006 | 0.0006 |
|  | GM pericalcarine L | -0.0302 | 0.0141 | 303 | -2.1360 | **0.0335** | -0.0580 | -0.0024 | 0.0014 | 0.0014 | 0.0003 | 0.0003 |
|  | GM pericalcarine R | -0.0425 | 0.0149 | 304 | -2.8549 | **0.0046** | -0.0718 | -0.0133 | 0.0015 | 0.0015 | 0.0003 | 0.0003 |
|  | NAWM cuneus L | -0.0355 | 0.0140 | 303 | -2.5362 | **0.0117** | -0.0630 | -0.0080 | 0.0016 | 0.0016 | 0.0003 | 0.0003 |
|  | NAWM cuneus R | -0.0019 | 0.0149 | 303 | -0.1298 | 0.8968 | -0.0313 | 0.0274 | 0.0016 | 0.0016 | 0.0003 | 0.0003 |
|  | NAWM lateral occipital L | -0.0691 | 0.0147 | 305 | -4.7133 | **0.0000** | -0.0979 | -0.0403 | 0.0060 | 0.0060 | 0.0007 | 0.0007 |
|  | NAWM lateral occipital R | -0.0459 | 0.0118 | 304 | -3.8868 | **0.0001** | -0.0691 | -0.0227 | 0.0062 | 0.0062 | 0.0008 | 0.0008 |
|  | NAWM lingual L | -0.1192 | 0.0153 | 304 | -7.7776 | **0.0000** | -0.1493 | -0.0891 | 0.0033 | 0.0033 | 0.0005 | 0.0005 |
|  | NAWM lingual R | -0.0813 | 0.0135 | 305 | -6.0041 | **0.0000** | -0.1078 | -0.0547 | 0.0035 | 0.0035 | 0.0006 | 0.0006 |
|  | NAWM pericalcarine L | -0.1622 | 0.0130 | 305 | -12.5033 | **0.0000** | -0.1876 | -0.1367 | 0.0020 | 0.0019 | 0.0004 | 0.0004 |
|  | NAWM pericalcarine R | -0.1386 | 0.0122 | 304 | -11.3367 | **0.0000** | -0.1626 | -0.1146 | 0.0020 | 0.0020 | 0.0004 | 0.0005 |
| Insula | GM insula L | -0.0466 | 0.0172 | 303 | -2.7129 | **0.0071** | -0.0802 | -0.0129 | 0.0045 | 0.0045 | 0.0004 | 0.0004 |
|  | GM insula R | -0.0408 | 0.0193 | 305 | -2.1147 | **0.0353** | -0.0787 | -0.0029 | 0.0045 | 0.0045 | 0.0004 | 0.0004 |
|  | NAWM insula L | -0.0368 | 0.0147 | 303 | -2.5082 | **0.0127** | -0.0656 | -0.0080 | 0.0061 | 0.0061 | 0.0005 | 0.0005 |
|  | NAWM insula R | -0.0178 | 0.0188 | 304 | -0.9494 | 0.3432 | -0.0547 | 0.0190 | 0.0060 | 0.0060 | 0.0005 | 0.0005 |
| B=standardised beta value, SE=standard error, df=degrees of freedom, CI=confidence interval for beta value, SD=standard deviation, w0=baseline, w1=1-year follow-up, L=left, R=right, GM=grey matter, NAWM=normal-appearing white matter, WML=white matter lesion, r-ICV=ratio of intracranial volume.  p-values surviving FDR correction (q<0.05) are highlighted in bold | | | | | | | | | | | | |
